# Longitudinal trajectories across the Command, Modifier, and Syntactic Phenotypes of language comprehension in over 6,000 autistic children

**DOI:** 10.64898/2025.12.19.25342690

**Authors:** Rohan Venkatesh, Anetka Nowakowski, Edward Khokhlovich, Andrey Vyshedskiy

## Abstract

Typically-developing children progress through three distinct language-comprehension phenotypes. 1) The Command Phenotype, emerging by age 2, is characterized by understanding single words and simple commands. 2) The Modifier Phenotype, observed around age 3, is characterized by understanding adjective–noun combinations. 3) The Syntactic Phenotype, reached by age 4, is characterized by understanding stories and complex syntactic structures. This study examined language-comprehension trajectories in autistic children using parent-submitted longitudinal assessments from 6,736 participants, with a mean observation period of 2.2 ± 1.3 years, spanning ages 1.5–22 years. Autistic children advanced through the same three phenotypes as neurotypical children but showed systematic differences. Increasing autism severity both reduced the likelihood of attaining higher-level phenotypes and lengthened the time required to reach them. The Command Phenotype was retained by 11%, 19%, and 39% of individuals with mild, moderate, and severe autism. Among individuals who advanced, median ages for acquiring the Modifier Phenotype were 3.7, 4.6, and 5.7 years for those with mild, moderate, and severe autism. For the Syntactic Phenotype, median ages were 4.8, 5.9, and 6.5 years across the same groups. These findings provide the first large-scale quantification of language-comprehension trajectories in autism and underscore the importance of early intervention.

## Introduction

The most compelling insights in biology emerge from dynamic processes. In particular, on multiple occasions researchers have studied longitudinal trajectories of language development in children with impairments ^1–9^. Collectively, the results of these studies indicate that language development during the preschool years is highly variable: for many children, early language difficulties eventually resolve ^10–12^, whereas those who continue to have poor language skills at the age of five are unlikely to catch up later ^13–15^.

Language acquisition is shaped by both immutable factors, such as genetics, and modifiable factors that can be influenced through interventions with the child or their family, including language therapy, increased book reading, shared play, reduced video viewing, and improved diet ^16–28^. The potential to enhance language development, coupled with evidence that such interventions are most effective in early childhood, has fueled strong interest in the early detection of language impairments and in predicting their developmental trajectory.

Our recent identification of three distinct, quantized language comprehension phenotypes offers a powerful framework for understanding language-acquisition trajectories. Drawing on analyses of tens of thousands of individuals with language impairments ^29–31^, we define these phenotypes as follows: 1) The Syntactic Phenotype is exhibited by most adults and by children aged four years and older ^32^. Individuals with this phenotype can comprehend sentences containing spatial prepositions, reversible word order, verb tenses, possessive pronouns, complex explanations, and fairytales (Table 1, Syntactic Mechanism). 2) Approximately 2% of adults, as well as children between the ages of three and four, exhibit the Modifier Phenotype. Their ability is limited to combinations of nouns and adjectives (e.g., they can select ‘*a large red straw*’ from a set of pencils, straws, and Lego pieces of different sizes and colors; Table 1, Modifier Mechanism). Adolescents and adults with the Modifier Phenotype typically have IQs between 50 and 70 and may be minimally to fully verbal. 3) About 1% of adults, and children younger than three years, exhibit the Command Phenotype, being restricted to simple commands (e.g., ‘eat banana’; Table 1, Command Mechanism). The Command Phenotype is commonly observed in adolescents and adults with profound autism; their IQ is typically below 50, and they are usually nonverbal, although any level of verbal ability can co-occur with any comprehension phenotype. These three language-comprehension phenotypes have proven remarkably stable across a wide range of factors, including gender, native language, verbal ability, parental education, and diagnostic categories (e.g., ASD, mild language delay, apraxia, Specific Language Impairment, Sensory Processing Disorder, Social Communication Disorder, Down Syndrome, ADHD) ^29–31^.

**Table 1.** Three language comprehension mechanisms—Syntactic, Modifier, and Command—have been identified in previous studies^29–31^. When one mechanism is acquired, the entire range of associated comprehension abilities is also gained. The Command-level abilities (Items 1 to 4) are acquired first. The Modifier-level abilities (Items 5 to 8) are attained next. The Syntactic-level abilities (Items 9 to 15) are acquired last. The language comprehension items are presented exactly as surveyed with parents in both current and earlier studies. Response options were: very true, somewhat true, and not true. Items 1 to 3 were assessed as part of the Expressive Language ATEC ^39^ subscale 1; the rest of items were a part of the MSEC subscale ^40^.

|  |  | Language comprehension items (verbatim) | Abbreviations used in dendrograms |
| --- | --- | --- | --- |
| Command Mechanism | 1 | Knows own name | Knows Name |
|  | 2 | Responds to 'No' or 'Stop' | No and Stop |
|  | 3 | Responds to praise | Resp. to Praise |
|  | 4 | Can follow some commands | Commands |
| Modifier Mechanism | 5 | Understands some simple modifiers (i.e., green apple vs. red apple or big apple vs. small apple) | Color or Size / Modifiers |
|  | 6 | Understands several modifiers in a sentence (i.e., small green apple) | Two Modifiers |
|  | 7 | Understands size (can select the largest/smallest object out of a collection of objects) | Size Superlatives |
|  | 8 | Understands NUMBERS (i.e., two apples vs. three apples) | Numbers |
| Syntactic Mechanism | 9 | Understands spatial prepositions (i.e., put the apple ON TOP of the box vs. INSIDE the box vs. BEHIND the box) | Sp. Prepositions |
|  | 10 | Understands verb tenses (i.e., I will eat an apple vs. I ate an apple) | Verb Tenses |
|  | 11 | Understands simple stories that are read aloud | Simple Stories |
|  | 12 | Understands elaborate fairytales that are read aloud (i.e., stories describing FANTASY creatures) | Elab. Fairytales |
|  | 13 | Understands possessive pronouns (i.e., your apple vs. her apple) | Poss. Pronouns |
|  | 14 | Understands the change in meaning when the order of words is changed (i.e., understands the difference between 'a cat ate a mouse' vs. 'a mouse ate a cat') | Flexible Syntax |
|  | 15 | Understands explanations about people, objects or situations beyond the immediate surroundings (e.g., "Mom is walking the dog," "The snow has turned to water"). | Explanations |

The Command, Modifier, and Syntactic Phenotypes were identified through data-driven clustering techniques that sorted linguistic abilities according to their co-occurrence ^29–31^. Each phenotype represents a coherent set of abilities that co-develop and decline together, consistent with distinct neural substrates. Notably, the distinction between the Modifier and Syntactic Mechanisms was unforeseen by traditional linguistic theory, and existing terminology—such as *propositional*, *grammatical*, *compositional*, and *Merge*—does not discriminate between them. A brief clarification of terminology is therefore warranted in order to demonstrate how existing frameworks fail to accommodate the Modifier Phenotype.

A *propositional* sentence is defined as a declarative statement that expresses a complete thought and can be evaluated as either *true* or *false*. In other words, it is a statement whose truth can be debated. Command sentences (e.g., “Bring the towel to the bathroom”) cannot be propositional, because they cannot be judged as true or false. However, both Modifier and Syntactic sentences can be *propositional*. For example, Modifier sentences such as “The sky is blue,” “The sky is red,” and “The sky is black” all meet the definition of propositional statements, as each can be evaluated for truth. The Syntactic sentence “The dog carries the cat” is also propositional. Since the comprehension of the former is mediated by the Modifier Mechanism and that of the latter by the Syntactic Mechanism, the term *propositional* fails to distinguish between these two mechanisms.

The term *compositional* refers to the principle that the meaning of a complex expression is determined by the meanings of its individual parts and the rules used to combine them. Both Modifier and Syntactic sentences can be *compositional*. For example, in the Modifier phrase “red apple,” the meaning is derived from the meanings of “red” and “apple” and how they are combined. Similarly, the Syntactic sentence “The dog rides the cat” is also *compositional.* Accordingly, the term *compositional* fails to differentiate between the Modifier and Syntactic Mechanisms.

The term *grammatical* has a similar issue. For instance, the definition of a “grammatical language” includes the use of inflections. Both Modifier and Syntactic sentences can use inflections. A sentence “This is the biggest apple” is deemed *grammatical* because the adjective “big” is inflected to form the superlative “biggest.” However, comprehension of superlatives is mediated by the Modifier Mechanism; an individual with only the Modifier Phenotype is still capable of understanding this sentence. As a result, the term *grammatical* fails to differentiate between the Modifier and Syntactic Mechanisms.

*Merge* is defined as an operation that combines two linguistic units (e.g., ‘green’, ‘dog’) to form a set (‘green dog’), which can then be further combined with additional linguistic units ^33,34^. The combination of an adjective and a noun to form of a set like ‘green dog’ is mediated by the Modifier Mechanism, whereas the combination of two nouns governed by a spatial preposition to form a set like ‘dog under table’ is mediated by the Syntactic Mechanism. Accordingly, the *Merge* operation is defined in a way that encompasses both the Modifier and Syntactic Mechanisms.

The absence of an established term to distinguish the Modifier Phenotype from the Syntactic Phenotype reveals a deeper conceptual blind spot within linguistic theory. Traditional frameworks correctly anticipated the contrast between the Syntactic Phenotype and the Command Phenotype, but they failed to envision a third, intermediate outcome; specifically, that a substantial number of individuals with fluent speech might acquire the Modifier Mechanism—supporting rich lexical, morphological, and local modification abilities—without ever acquiring the Syntactic Mechanism. In other words, linguistic theory never contemplated the possibility of a stable, lifelong Modifier Phenotype. The reasons causing some children to plateau at this stage remain an open question.

The identification of the three comprehension phenotypes holds broad clinical relevance. Age-normalized phenotype assessments ^32^ can facilitate earlier identification of comprehension difficulties and more precise monitoring of children’s developmental progress. In turn, this may enable earlier and more targeted interventions, ultimately helping children to attain higher levels of language comprehension. Furthermore, knowledge of a child’s comprehension phenotype can inform the design of individualized therapy programs. E. g., children with the Command Phenotype may benefit most from exercises emphasizing color, size, and number modifiers, whereas tasks targeting Syntactic Phenotype-level comprehension may be excessively demanding.

While our previous analyses of comprehension phenotypes relied on cross-sectional designs, the present study sought to characterize the longitudinal trajectories of children as they advance through the three phenotypes. We hypothesized that autistic children progress through the same three discrete language-comprehension phenotypes observed in neurotypical development (Command → Modifier → Syntactic) but do so at a substantially slower rate. A secondary objective was to assess the extent to which autistic participants exhibit stagnation at the lower-level phenotypes. Finally, we aimed to identify the oldest ages at which upward transitions occurred, thereby providing insight into the outer limits of the critical period for acquiring the Modifier and Syntactic Phenotypes.

## Methods

### Study Participants

Participants were children using a language therapy app that was made freely available at all three major app stores (Apple App Store, Google Play Store, and Amazon App Store) in September 2015 ^22,35–38^. The app provides various structured language comprehension therapy exercises and is primarily used by parents of children with language impairments. Once the app was downloaded, caregivers were asked to register and to provide demographic details, including the child’s diagnosis and age. Caregivers consented to pseudonymized data analysis and completed a 133-item questionnaire (77-item Autism Treatment Evaluation Checklist ^39^, Supplementary Tables 1 to 4; 20-item Mental Synthesis Evaluation Checklist (MSEC) ^40^, Supplementary Table 5; 10-item screen time checklist ^18^; 25-item diet checklist ^19^; and 1-item parent education survey) approximately every 4.8 months.

All fifteen available language comprehension items from the 133-item questionnaire were included in the cluster analysis as in previously published articles ^29–31^ (Table 1). Answer choices were as follows: very true (0 points), somewhat true (1 point), and not true (2 points). A lower score indicates better language comprehension ability.

The inclusion criteria for this study remained consistent with those of the previous studies ^29–31^: absence of seizures (which commonly result in intermittent, unstable language comprehension deficits ^41^) and absence of serious and moderate sleep problems (which are also associated with intermittent, unstable language comprehension deficits ^42^). Previous studies focused on individuals aged 4–22 years. To capture phenotypic transitions occurring earlier in development, we extended the age range in this study to 1.5–22 years. The minimum number of assessments was set at four, as required by R’s smooth.spline() function for a stable cubic smoothing spline fit. The study was limited to individuals diagnosed with autism spectrum disorder (ASD) and typically developing children. Autism level (mild/Level 1, moderate/Level 2, or severe/Level 3) was reported by caregivers. Pervasive Developmental Disorder and Asperger Syndrome were combined with mild autism for analysis as recommended by DSM-5 ^43^. A good reliability of such parent-reported diagnosis has been previously demonstrated ^44^.

Thus, the study included a total of 6,736 participants, the average age at baseline was 4.8±2.6 years (range of 1.5 to 20.8 years), 79% participants were males. Table 2 reports participants’ demographics in each diagnostic group as communicated by caregivers. The education level of participants’ parents was the following: 91.8% with at least a high school diploma, 71.4% with at least college education, 39.2% with at least a master’s, and 6.9% with a doctorate. All caregivers consented to pseudonymized data analysis and publication of results. The study was conducted in compliance with the Declaration of Helsinki ^45^. The study protocol was approved by the Biomedical Research Alliance of New York (BRANY) LLC Institutional Review Board (IRB). The data was accessed on July 9, 2025.

**Table 2.** Participants’ demographics.

|  | **Number of**<br>**Participants** | **Percent of**<br>**Total** | **Age at baseline,**<br>**Mean(SD)** | **Percent**<br>**Males** |
| --- | --- | --- | --- | --- |
| **Neurotypical** | 322 | 4.8 | 2.9(1.1) | 59 |
| **Mild ASD** | 3091 | 45.9 | 4.4(2.2) | 79 |
| **Moderate ASD** | 2262 | 33.6 | 5(2.5) | 79 |
| **Severe ASD** | 1061 | 15.8 | 6.1(3.5) | 83 |
| **Total** | 6736 | 100 | 4.8(2.6) | 79 |

### Statistics and Reproducibility

Unsupervised Hierarchical Cluster Analysis (UHCA) was performed using Ward’s agglomeration method with a Euclidean distance metric. The clustering was entirely data-driven, without any pre-specified design or hypothesis. A two-dimensional heatmap was generated using the R package “pheatmap” ^46^. This heatmap analysis was restricted to children aged 4 to 22 years, consistent with prior studies ^29–31^.

R’s smooth.spline() function was employed to estimate continuous developmental trajectories. This procedure fits a cubic smoothing spline by minimizing a penalized least-squares criterion that balances fidelity to the observed data with curvature of the fitted curve. The smoothing parameter is selected automatically, yielding a trajectory that captures broad developmental trends while suppressing noise in individual measurements.

Code and data can be downloaded from https://doi.org/10.6084/m9.figshare.30801752.

## Results

### Clustering analysis of 15 language comprehension abilities

Caregivers assessed 15 language comprehension abilities (Table 1) over time. Whereas prior analyses relied on cross-sectional designs and therefore used only the final available assessment for each participant ^29–31^, the present study investigated longitudinal development in language comprehension and consequently included all assessments for every participant. As in previous analyses, the first step was to examine patterns of co-occurrence among the 15 abilities, using unsupervised hierarchical cluster analysis (UHCA), a data-driven method that groups items based on their similarity. This technique produces dendrograms, which visually represent the hierarchical relationships between clusters of items. Abilities that frequently co-occur are positioned in closer proximity, while those that co-occur less often appear farther apart.

Figure 1A depicts the dendrogram. The height of the branches indicates the distance between clusters. A larger distance corresponds to greater dissimilarity between the clusters. Previous studies identified three clusters that were stable across different evaluation methods, age groups, time points, genders, native languages, and levels of parental education ^29–31^. The first cluster included *knowing the name, responding to ‘No’ or ‘Stop’, responding to praise*, and *following some commands* (items 1 to 4 in Table 1) and was termed the Command Mechanism. The second cluster included *understanding color and size modifiers, several modifiers in a sentence, size superlatives*, and *numbers* (items 5 to 8 in Table 1) and was termed the Modifier Mechanism. The third cluster included *understanding of spatial prepositions, verb tenses, flexible syntax, possessive pronouns, explanations about people and situations, simple stories*, and *elaborate fairytales* (items 9 to 15 in Table 1) and was termed the Syntactic Mechanism. The analysis presented in Figure 1A identified the same three clusters with inter-cluster distances that were significantly larger than the distances between subclusters. Principal component analysis (PCA) (Figure 1B) also showed a clear separation between these three clusters: Command, Modifier and Syntactic.

**Figure 1.**
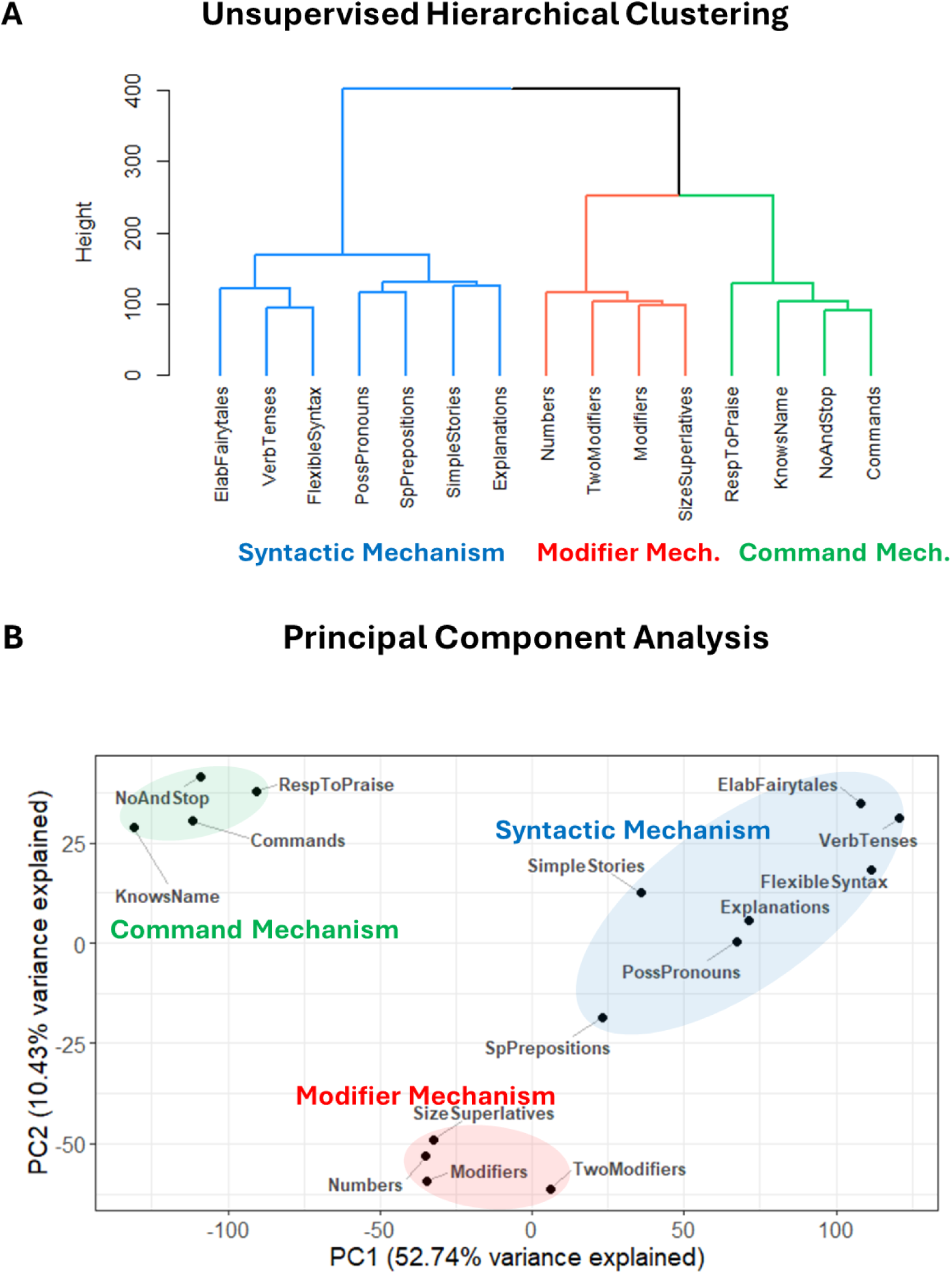
Clustering analysis of 15 comprehension items. (A) A dendrogram representing the unsupervised hierarchical clustering analysis (UHCA) of language comprehension abilities. (B) Principal component analysis (PCA) plot, where ovals highlight clusters identified by UHCA. The PCA reveals a distinct separation among Command, Modifier and Syntactic Mechanisms. Principal component 1 accounts for 47% of the variance in the data. Principal component 2 accounts for 11.1% of the variance in the data.

As a control we calculated UHCA and PCA of the 15 comprehension abilities along with the “hyperactivity” (Figures S1), “bed-wetting” (Figure S2), and “demands sameness” (Figure S3) items. These items are not related to language and therefore should cluster into their own group. As expected, both UHCA and PCA clustered these items into their own group at a significant distance from the Command, Modifier, and Syntactic clusters, validating both clustering techniques.

### Language-comprehension phenotypes in participants

Previous studies have employed UHCA to identify distinct language-comprehension phenotypes of participants ^29–31^. The principles underlying participant clustering are identical to those used for clustering abilities: participants with similar patterns of abilities are automatically organized into hierarchical dendrograms. Previous studies identified three distinct phenotypes: 1) the Command Phenotype – participants who acquired only the Command Mechanism; 2) the Modifier Phenotype – participants who acquired both the Command and Modifier Mechanisms; and 3) the Syntactic Phenotype – participants who acquired the Command, Modifier, and Syntactic Mechanisms.

The heatmap in Figure 2 relates participant clusters to comprehension mechanisms. The three mechanism clusters (Command, Modifier, and Syntactic) are shown as rows (the dendrogram from Figure 1A representing comprehension mechanisms is shown vertically on the left in Figure 2) and the participants are shown as columns (the dendrogram representing participants is shown horizontally on the top). *Blue* indicates the presence of a linguistic ability (parent’s response=*very true*); *white-yellow* indicates an intermittent presence of a linguistic ability (parent’s response=*somewhat true*); and *red* indicates the complete lack of a linguistic ability (parent’s response=*not true*). The rightmost cluster of participants (marked “Syntactic Phenotype”) shows the predominant blue color (representing good skills) across all abilities indicating that these participants acquired the Command, Modifier, and Syntactic Mechanisms. The leftmost cluster of participants (marked “Command Phenotype”) shows the predominant blue color only across the Command Mechanism items and red colors across Syntactic and Modifier Mechanisms items, indicating that these individuals only acquired the Command Mechanism. The center cluster of participants (marked “Modifier Phenotype”) shows the predominant blue color only across Command and Modifier Mechanisms items and white to red colors across Syntactic Mechanism items, indicating that these individuals acquired the Command and Modifier Mechanisms.

**Figure 2.**
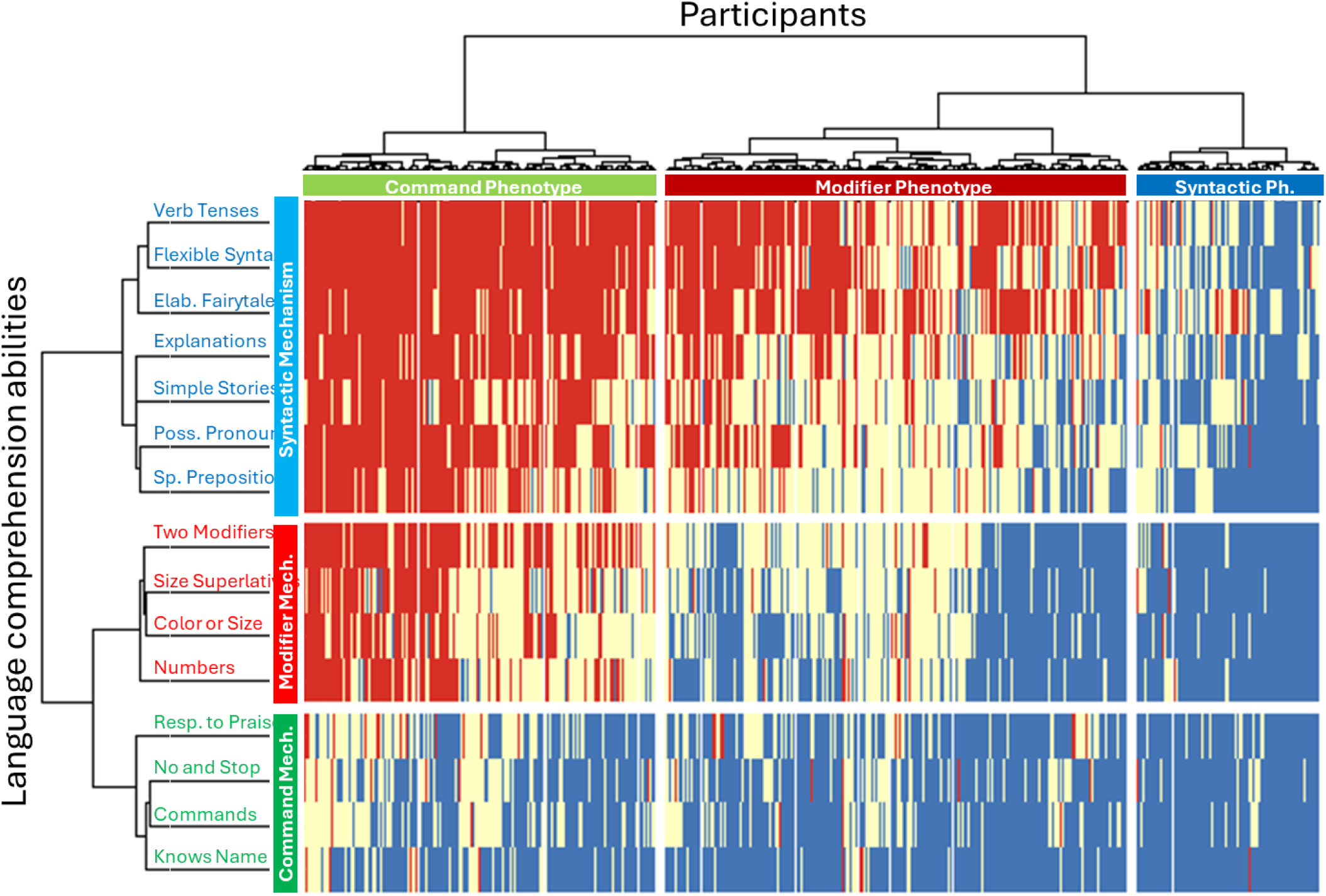
Two-dimensional heatmap relating participants to their language comprehension abilities. The 15 language comprehension abilities are shown as rows. The dendrogram representing language comprehension abilities is shown on the left. Participants are shown as columns. The dendrogram representing participants is shown on the top. Blue color indicates the presence of a linguistic ability (the “very true” answer), red indicates the lack of a linguistic ability (the “not true” answer), and white-yellow indicates the “somewhat true” answer.

The PCA plot corroborates the findings from the UHCA analysis. As shown in Figure 3, participants identified by the UHCA as having the Command Phenotype are depicted in green, those with the Modifier Phenotype in red, and those with the Syntactic Phenotype in blue. Ellipses show 95% confidence intervals for each phenotype. On the plot, participants with the Command Phenotype cluster toward the right (PC1>=1.4), those with the Modifier Phenotype occupy the center (−2<=PC1<1.4), and those with the Syntactic Phenotype are concentrated on the left (PC1<-2). Accordingly, as children improved their phenotypes their trajectory was expected to follow from right to left of the PCA plot.

**Figure 3.**
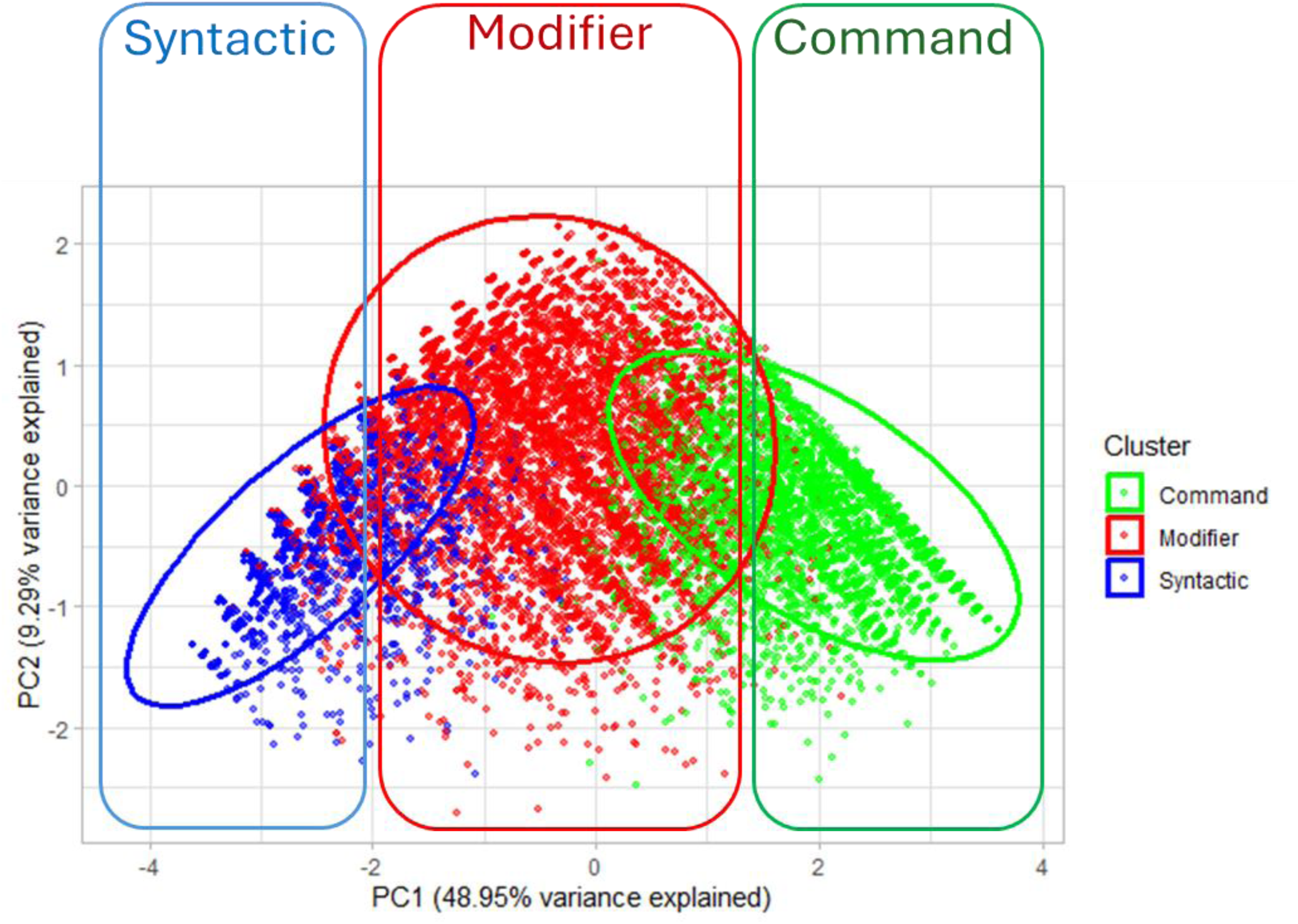
Distribution of comprehension phenotypes in PCA space. Participants identified by the UHCA as having the Command Phenotype are shown in green, those with the Modifier Phenotype in red, and those with the Syntactic Phenotype in blue. Ellipses represent 95% confidence intervals for each phenotype. Participants with the Command Phenotype cluster toward the right (PC1 ≥ 1.4), those with the Modifier Phenotype occupy the central region (−2 ≤ PC1 < 1.4), and those with the Syntactic Phenotype cluster toward the left (PC1 < −2). Improvements in phenotype correspond to a right-to-left trajectory along the PC1 axis.

### Longitudinal changes in comprehension phenotype

The progression of all participants across comprehension phenotypes as a function of age is visualized in the PCA plot (Figure S4). Each trajectory begins at the age of the participant’s first assessment and ends at the age of their last assessment, connected by a spline function computed from all available observations. As shown in Figure 3, the right side of the PCA plot corresponds to the Command Phenotype, the center to the Modifier Phenotype, and the left to the Syntactic Phenotype. The movie in Figure S4 indicates that most participants start on the right side of the PCA plot and gradually shift leftward over time, reflecting a progression from the Command to the Modifier and, ultimately, to the Syntactic Phenotype.

At the final reported observation, 25% of participants had attained the Syntactic Phenotype, 58% had reached the Modifier Phenotype, and 17% remained at the Command Phenotype (Table 3). Table 4 presents median age at the final assessment for each phenotype and each diagnostic group. The likelihood of children remaining at the Command Phenotype increased with ASD severity. At the final assessment, 39% of children with severe ASD remained at this level, compared with 19% of those with moderate ASD, 11% with mild ASD, and only 2% of neurotypical children. The likelihood of children reaching the Syntactic Phenotype decreased with ASD severity. At the final assessment, only 7% of children with severe ASD attained this level, compared with 15% with moderate ASD, 33% with mild ASD, and 73% of neurotypical children.

**Table 3.** Proportion of participants exhibiting the Command, Modifier, and Syntactic phenotypes at their final assessment, stratified by diagnostic group.

|  | **Neurotypical** | **Mild ASD** | **Moderate ASD** | **Severe ASD** | **Any diagnosis** |
| --- | --- | --- | --- | --- | --- |
| **Syntactic Phenotype** | 73% | 33% | 15% | 7% | 25% |
| **Modifier Phenotype** | 25% | 56% | 66% | 55% | 58% |
| **Command Phenotype** | 2% | 11% | 19% | 39% | 17% |
| **Total** | 100% | 100% | 100% | 100% | 100% |

**Table 4.** Median age and interquartile range (IQR) at the final assessment for each phenotype, stratified by diagnostic group (years). The age at the final assessment increases systematically with ASD severity across phenotypes.

|  | **Neurotypical** | **Mild ASD** | **Moderate ASD** | **Severe ASD** | **Any diagnosis** |
| --- | --- | --- | --- | --- | --- |
| **Syntactic Phenotype** | 4.6(3.8-5.5) | 5.8(4.8-7.1) | 7.1(5.7-8.6) | 8.7(6.5-10.2) | 5.9(4.8-7.4) |
| **Modifier Phenotype** | 3.6(3.1-4.4) | 5.5(4.4-7.1) | 6.7(5.3-8.6) | 7.8(5.8-10.1) | 6.0(4.7-8.0) |
| **Command Phenotype** | 2.6(2.3-3.4) | 4.3(3.3-6.0) | 5.3(4.1-7.3) | 6.8(5.0-9.6) | 5.2(3.9-7.4) |

### Differences in age of acquisition of comprehension phenotypes across diagnostic groups

In the movie (Figure S4), neurotypical participants are shown in black, and individuals with mild, moderate, and severe ASD in blue, red, and green, respectively. The visualization reveals four temporal waves moving from right to left across the PCA plot, each wave reflecting the developmental transition from Command → Modifier → Syntactic Phenotype. The first wave, dominated by neurotypical participants, peaks between 1.5 to 3.5 years of age. It is followed by successive waves of children with mild ASD peaking between 2 to 5 years of age, those with moderate ASD peaking between 2.5 to 7 years of age, and those with severe ASD peaking between 3 to 8 years of age. This systematic delay shows that as ASD severity increases, the acquisition of advanced comprehension phenotypes becomes progressively more delayed.

A similar developmental sequence is evident in the individual participant examples presented in Figure 4. The neurotypical participant in Figure 4A acquired the Modifier Phenotype at approximately 3.1 years of age and the Syntactic Phenotype at 4.5 years. The neurotypical participant in Figure 4B reached the Modifier Phenotype at about 2.5 years and the Syntactic Phenotype at 3.6 years, while the neurotypical participant in Figure 4C reached the Modifier Phenotype at approximately 3.0 years and the Syntactic Phenotype at 4.6 years.

**Figure 4.**
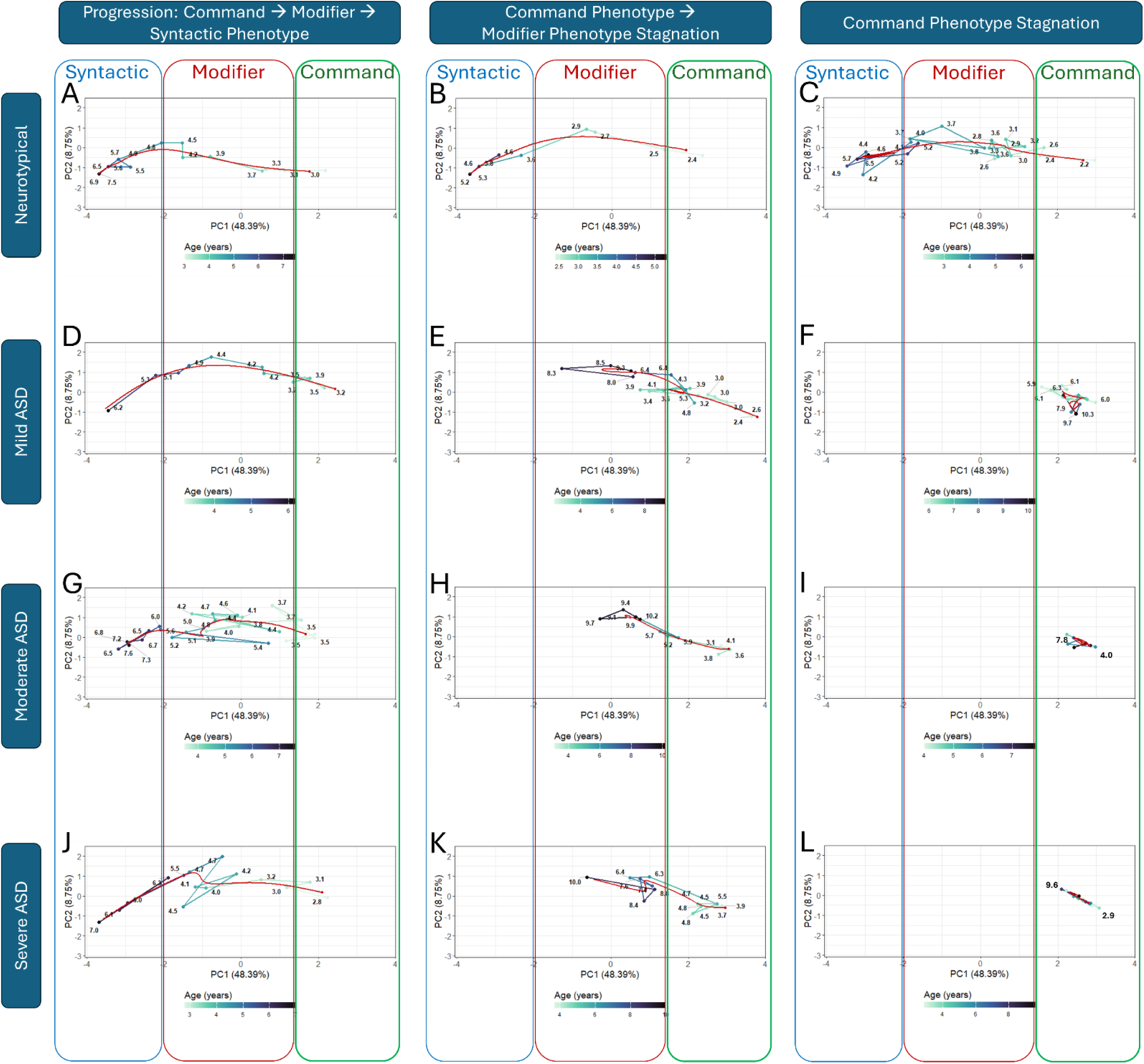
Longitudinal phenotype trajectories on the PCA plot for 12 representative participants. Each circle represents an assessment, with the adjacent number reporting the child’s age at that assessment. The color of the lines connecting successive assessments indicates age. Improvements in phenotype correspond to a right-to-left movement along the PC1 axis. The red smooth line shows a piecewise polynomial fit (spline). (A) Neurotypical participant acquired the Modifier Phenotype at ∼3.1 years and the Syntactic Phenotype at 4.5 years. (B) Neurotypical participant reached the Modifier Phenotype at ∼2.5 years and the Syntactic Phenotype at 3.6 years. (C) Neurotypical participant acquired the Modifier Phenotype at ∼3.0 years and the Syntactic Phenotype at 4.6 years. (D) Participant with mild ASD acquired the Modifier Phenotype at ∼3.5 years and the Syntactic Phenotype at 5.2 years. (E) Participant with mild ASD reached the Modifier Phenotype at ∼3.5 years and remained at this level through at least 9.3 years (final assessment). (F) Participant with mild ASD remained at the Command Phenotype for at least 6.4 years (from 5.9 to 10.3 years, last assessment). (G) Participant attained the Modifier Phenotype at ∼3.5 years and the Syntactic Phenotype at 6 years. (H) Participant reached the Modifier Phenotype at ∼5.2 years and remained at this level through at least 10.2 years (last assessment). (I) Participant with moderate ASD remained at the Command Phenotype for at least 3.8 years (from 4.0 to 7.8 years, last assessment). (J) Participant acquired the Modifier Phenotype at 3.2 years and the Syntactic Phenotype at 6.3 years. (K) Participant reached the Modifier Phenotype at ∼6.3 years and remained at this level through at least 10 years (last assessment). (L) Participant with severe ASD remained at the Command Phenotype for at least 6.7 years (from 2.9 to 9.6 years, last assessment).

Participants with mild ASD show slightly delayed trajectories. The participant in Figure 4D acquired the Modifier Phenotype at around 3.5 years and the Syntactic Phenotype at 5.2 years. The participant in Figure 4E also reached the Modifier Phenotype at approximately 3.5 years, but remained at this developmental level through at least 9.3 years of age, the time of the final assessment.

Participants with moderate ASD exhibited further delays. The participant in Figure 4G attained the Modifier Phenotype at around 3.5 years and the Syntactic Phenotype at 6 years. The participant in Figure 4H reached the Modifier Phenotype at approximately 5.2 years and remained at this level through at least 10.2 years, the last recorded assessment.

Finally, participants with severe ASD showed the most pronounced delay in developmental progression. The participant in Figure 4J acquired the Modifier Phenotype at 3.2 years and the Syntactic Phenotype at 6.3 years. The participant in Figure 4K reached the Modifier Phenotype at approximately 6.3 years and remained at this level through at least 10 years of age, the last recorded assessment.

As shown in Figure 5A, the frequency distributions of age at which participants reached the Modifier Phenotype differed systematically across groups. Median ages of acquisition increased with ASD severity, from 2.5 years in neurotypical participants to 3.7, 4.6, and 5.7 years in the mild, moderate, and severe ASD groups, respectively (Figure 5B).

**Figure 5.**
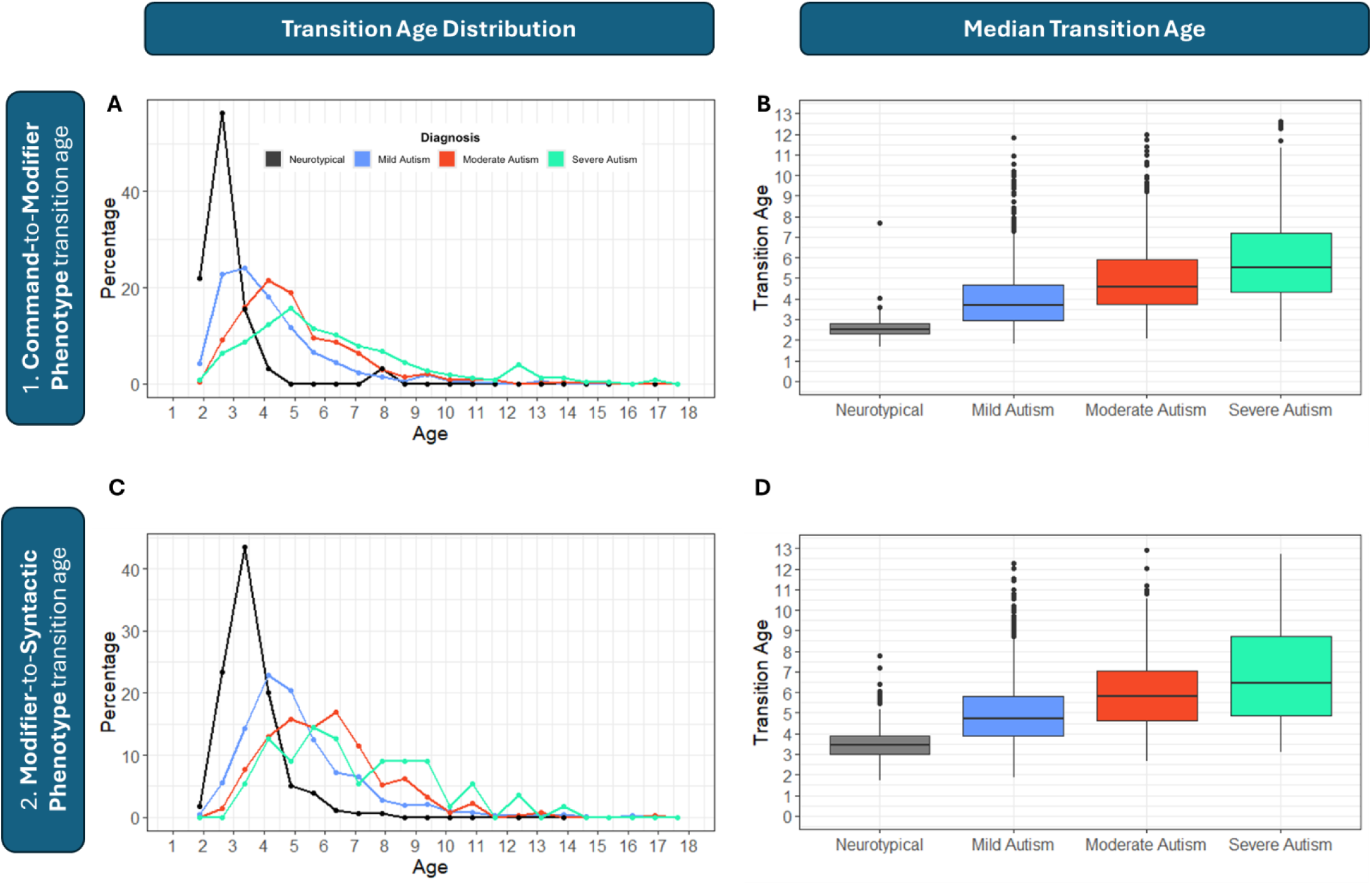
Age-of-acquisition of each language-comprehension phenotype. (A) Distribution of ages when children transition from the Command to the Modifier Phenotype. (B) Box plot indicates the median age and quartiles for the transition from the Command to the Modifier Phenotype. All pairwise group comparisons are statistically significant (Wilcoxon test, *p* < 0.0001). (C) Distribution of ages when children transition from the Modifier to the Syntactic Phenotype. (D) Box plot indicates the median age and quartiles for the transition from the Modifier to the Syntactic Phenotype. All pairwise group comparisons are statistically significant (Wilcoxon test, *p* < 0.0001; for the comparison between moderate and severe ASD groups *p* = 0.02).

A comparable pattern was observed for the Syntactic Phenotype (Figure 5C), with median ages of acquisition of 3.5, 4.8, 5.9, and 6.5 years for the neurotypical, mild, moderate, and severe ASD groups, respectively (Figure 5D).

### Stagnation at the Command and Modifier Phenotypes

Whereas neurotypical participants tended to progress quickly through the Command and Modifier Phenotypes, participants with ASD often exhibited multi-year periods during which their assessments clustered within the Command or Modifier regions of the PCA plot. Several examples of stagnation at the Command Phenotype are shown in Figure 4. The participant with mild ASD (Figure 4F) remained at the Command level for at least 6.4 years (from 5.9 to 10.3 years of age, last assessment). The participant with moderate ASD (Figure 4I) showed stagnation at the Command level for at least 3.8 years (from 4.0 to 7.8 years, last assessment), and the participant with severe ASD (Figure 4L) remained at the Command level for at least 6.7 years (from 2.9 to 9.6 years, last assessment).

Prolonged stagnation was also observed at the Modifier Phenotype. The participant with mild ASD (Figure 4E) remained at the Modifier level for at least 5.8 years (from approximately 3.5 to 9.3 years of age, last assessment). The participant with moderate ASD (Figure 4H) showed stagnation at the Modifier level for at least 5 years (from 5.2 to 10.2 years, last assessment), and the participant with severe ASD (Figure 4K) remained at the Modifier level for at least 3.7 years (from approximately 6.3 to 10 years, last assessment).

Group-level analyses confirmed the trends observed in individual trajectories. Neurotypical participants progressed through the Command Phenotype more quickly, only spending a median of 0.3 years in this stage, compared with 0.6 years for individuals with mild ASD, 1.0 year for moderate ASD, and 1.3 years for severe ASD (Table 5). A similar pattern was evident in the Modifier Phenotype, where neurotypical individuals spent a median of 0.9 years, versus 1.2 years for mild ASD, 1.6 years for moderate ASD, and 1.4 years for severe ASD.

**Table 5.** Median years and interquartile range (IQR) participants spent in each phenotype, presented separately for each diagnostic group. The time spent in the Modifier and Command Phenotypes increases systematically with ASD severity across phenotypes.

|  | **Neurotypical** | **Mild ASD** | **Moderate ASD** | **Severe ASD** | **Any diagnosis** |
| --- | --- | --- | --- | --- | --- |
| **Syntactic Phenotype** | 1.0(0.5-1.6) | 0.9(0.4-1.6) | 0.9(0.4-1.8) | 0.8(0.4-1.4) | 0.9(0.4-1.6) |
| **Modifier Phenotype** | 0.9(0.5-1.4) | 1.2(0.7-2.0) | 1.6(0.9-2.5) | 1.4(0.7-2.4) | 1.3(0.8-2.2) |
| **Command Phenotype** | 0.3(0.1-0.6) | 0.6(0.5-1.2) | 1.0(0.4-1.7) | 1.3(0.7-2.3) | 0.9(0.4-1.7) |

The number of participants transitioning to a higher-level phenotype in any diagnostic category decreases dramatically after 5 years of age and nearly completely ceases after 10 years of age (Figures 4S, 5S).

## Discussion

This longitudinal analysis reveals that autistic children progress through the same three discrete language-comprehension phenotypes previously tracked over time in neurotypical development (Command → Modifier → Syntactic) ^32^, but that the maximum level and timing of their acquisition vary systematically with autism severity. The data demonstrates that the proportion of individuals reaching the advanced phenotypes decreases significantly with ASD severity, and that the time of acquisition is also significantly delayed. These findings provide the first large-scale quantification of developmental trajectories in comprehension phenotypes among autistic children.

### Impact of ASD severity on delayed acquisition of higher-level language phenotypes

Unsupervised hierarchical clustering analysis of participants (Figure 2) enabled us to map the three language phenotypes onto the principal component analysis (PCA) plot: the Command Phenotype (Figure 3, right), the Modifier Phenotype (center), and the Syntactic Phenotype (left). As children age, they advance through these phenotypes, a progression that can be followed by their trajectories moving from right to left on the PCA plot. This developmental “movie” is visualized in Figure S4, and individual examples are shown in Figure 4.

Figure 5 and supporting case examples reveal a clear, severity-dependent gradient of the age of higher-level phenotype acquisition. Median age at acquisition of the Modifier Phenotype increased progressively from 2.5 years in neurotypical children to 3.7 years (mild ASD), 4.6 years (moderate ASD), and 5.7 years (severe ASD). A parallel gradient was observed for the Syntactic Phenotype: 3.5 years (neurotypical), 4.8 years (mild ASD), 5.9 years (moderate ASD), and 6.5 years (severe ASD). These findings demonstrate that ASD severity exerts a strong, dose-dependent effect on the pace of language-comprehension development.

Importantly, the same ordered sequence of phenotypes is preserved across all groups, indicating that the underlying developmental architecture remains intact, though it progresses more slowly in children with greater ASD severity. These results align with prior cross-sectional findings ^29,30^ suggesting that autistic children are not atypical in *what* they learn, but rather in *when* and *how quickly* they acquire each linguistic mechanism.

The observed delays may stem from multiple, nonexclusive sources. Reduced social attention and communicative reciprocity could limit linguistic input, while atypical neural connectivity within fronto-parietal and fronto-temporal circuits may constrain the integration of syntactic and semantic information. Alternatively, motivational and sensory factors may impede participation in language-rich interactions critical for progression beyond the Command Phenotype. Future neuroimaging and intervention studies could help clarify whether these delays reflect intrinsic neural timing differences, environmental factors, or a combination of both.

### Stagnation at inferior phenotype levels

Approximately 17% of participants remained at the Command Phenotype at their final assessment at the median age of 5.2 years (see individual examples in Figures 4F, I, L), and another 58% remained at the Modifier Phenotype at their final assessment at the median age of 6.0 years (see examples in Figures 4E, H, K), consistent with our previous findings ^29–31^.

The proportion of individuals reaching the Syntactic Phenotype decreased significantly with increasing ASD severity, whereas the proportion remaining at the Command Phenotype increased with ASD severity (Table 3). Because we had no control over when parents submitted the final assessment, it is possible that some children classified within the Command or Modifier Phenotypes could have progressed to the Syntactic Phenotype after parental reporting ceased. However, the likelihood of such unobserved progression decreases both with age and duration of stagnation at a lower-level phenotype.

At the final assessment, only 2% of neurotypical participants classified within the Command Phenotype (Table 3). Their young median age of 2.6 years (Table 4) and short median duration in this phenotype, 0.3 years (Table 5), suggest that most would be expected to progress to the Modifier and subsequently to the Syntactic Phenotype. In contrast, 39% of participants with severe ASD exhibited the Command Phenotype at the final assessment. Their older median age of 6.8 years and longer median duration in this phenotype, 1.3 years, make such progression less likely.

A similar pattern was observed for the Modifier Phenotype. At the final assessment, only 25% of neurotypical participants remained at this level. Their young median age of 3.6 years and short median duration in this phenotype, 0.9 years, indicate that most would be expected to progress to the Syntactic Phenotype. In contrast, 55% of participants with severe ASD remained at the Modifier Phenotype. Their older median age of 7.8 years and longer median duration in this phenotype, 1.4 years, do not suggest a strong likelihood of further progression.

### Phenotype transitions cease at the onset of puberty

A notable finding is that upward phenotype transitions were rarely observed after approximately 10 years of age (Figures 5A and 5C; also see the movie in Figure 4S and Figure 5S). This apparent age boundary applies to both Modifier-to-Syntactic and Command-to-Modifier transitions, suggesting that the window for acquiring higher-level comprehension phenotypes may close around the onset of puberty. This boundary is consistent with prior evidence from linguistically-deprived individuals. For example, cases such as Genie ^47–49^ and studies of homesigners—deaf children born to hearing parents who use Command-level gestures (homesign or kitchensign) instead of a Syntactic-level formal sign language ^49–57^—demonstrate that children who are not involved in syntactic conversations before puberty are unable to acquire the Syntactic Phenotype after the onset of puberty.

Collectively, our results indicate that there are two major factors limiting the development of autistic children: 1) A strict temporal boundary near the onset of puberty and 2) A slower language-learning-rate. Recognizing these constraints has important implications for intervention, underscoring the need for therapies targeting syntactic comprehension to be implemented as early as possible in order to maximize their potential benefits.

To our knowledge, this is the largest ASD epidemiological study to provide clear evidence for both the lack of acquisition of higher-level language phenotypes after puberty and a slower language-comprehension learning-rate.

### Critical period for syntactic language acquisition

Critical (or sensitive) periods are defined as developmental windows during which neural circuits underlying specific behaviors or functions undergo experience-dependent plasticity, subsequently stabilize, and become resistant to perturbation ^58^. Although the existence of a critical period for a first syntactic language acquisition is broadly accepted, there is no consensus on its operational definition in measurable terms ^59^. A central unresolved question is whether this period is terminated by a uniform, hard maturational boundary at the onset of puberty, which would apply equally across neurotypical individuals and those with ASD, or whether its effective duration is instead governed by individual differences in the trajectory of learning-rate, resulting in a markedly shorter window in more severe ASD ^60,61^. Direct experimental investigation is inherently limited since full human language has no animal homologue, and human studies are restricted to rare and uncontrolled cases (e.g., feral children and late-exposed deaf homesigners) for obvious ethical reasons. The present epidemiological approach, leveraging large-scale longitudinal data across the full spectrum of ASD severity, offers a unique opportunity to adjudicate between these competing conceptualizations of the critical period for first language acquisition.

In a prior study, we characterized the trajectories of syntactic-language-comprehension-learning-rate in neurotypical and autistic children ^62^. We demonstrated that the rate of syntactic acquisition follows the canonical pattern of many biological growth processes (e.g., brain volume): an initially high and constant rate followed by exponential decay (Figure 6A). Early in development, autistic children exhibited learning-rates similar in amplitude to those of their neurotypical peers. In neurotypical children, however, this elevated rate was sustained until at least 7 years of age, whereas in autistic individuals, it began to decline exponentially substantially earlier—at approximately 3.2, 2.0, and 1.3 years in mild, moderate, and severe ASD, respectively (the “critical-slowing-down-age” ^62^; see Figure 6A). How do the present findings relate to these observations of earlier critical-slowing-down-age in autism?

**Figure 6.**
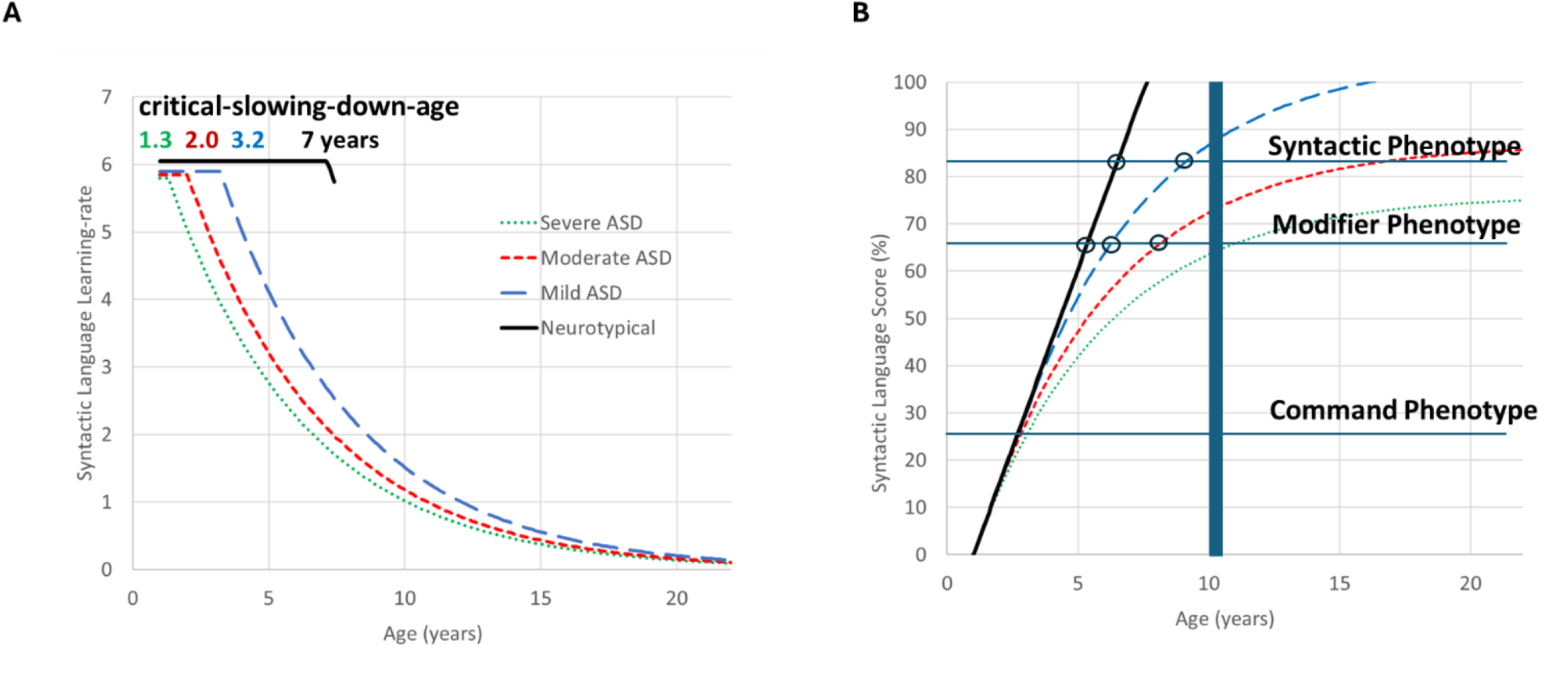
Differences in critical-slowing-down-ages determine syntactic-language “growth” curves. Adapted from our previously published work (Vyshedskiy et al. ^62^), which characterized trajectories of syntactic-language-comprehension-learning-rate in neurotypical and autistic children. (A) Early in development, autistic children exhibited learning-rates similar in amplitude to those of their neurotypical peers. In neurotypical children, however, this elevated rate was sustained until at least 7 years of age, whereas in autistic individuals, it began to exponentially decline at substantially earlier times, specifically, at approximately 3.2, 2.0, and 1.3 years in mild, moderate, and severe ASD, respectively (the “critical-slowing-down-age”) ^62^. (B) The syntactic-language-comprehension-learning-rate in Figure 6A can be translated into an absolute syntactic-language score by computing the area under the learning-rate curve. All four growth curves, corresponding to neurotypical, mild, moderate, and severe ASD, nearly overlap at the start. However, since the critical-slowing–down occurs earlier in ASD, the curves diverge over time, resulting in progressively lower ultimate levels of syntactic-language comprehension with increasing ASD severity. The three horizontal lines indicate the approximate language scores associated with the Command, Modifier, and Syntactic Phenotypes. The thick vertical line indicates the hard boundary at the onset of puberty.

The syntactic-language-comprehension-learning-rate in Figure 6A can be translated into an absolute syntactic-language score by computing the area under the learning-rate curve, as illustrated in Figure 6B. All four growth curves—corresponding to neurotypical, mild, moderate, and severe ASD— nearly overlap at the very start. However, since the critical-slowing–down occurs earlier in ASD, the curves diverge over time, resulting in progressively lower ultimate levels of syntactic-language comprehension with increasing ASD severity.

In Figure 6B, the three horizontal lines indicate the approximate language scores associated with the Command, Modifier, and Syntactic Phenotypes. Figure 6B predicts that by age 10 (the hard boundary at the onset of puberty indicated by the thick vertical line), most children with severe ASD will not reach the Modifier Phenotype; whereas most children with moderate ASD will reach the Modifier but not the Syntactic Phenotype; and children with mild ASD, as well as neurotypical children, will reach the Syntactic Phenotype.

Moreover, Figure 6B predicts that the age-of-acquisition for each phenotype (indicated by open circles) is progressively delayed in ASD in proportion to severity. Thus, the two main observations in this study align closely with our prior measurements of the critical-slowing-down-ages across diagnostic groups. In other words, the critical period for first-language acquisition cannot be reduced to a single number; rather, it reflects the combined effects of both earlier critical-slowing-down-ages in more severe ASD and the uniform hard boundary at the onset of puberty.

Figure 6B also illustrates why autistic children may be more sensitive to reduced linguistic input in early childhood compared to neurotypical children. The growth curves represent development under average conditions of language engagement—such as syntactic conversations and narrative input (e.g., fairy tales read aloud). When children experience lower-than-average engagement at home—for example, when interactive language is replaced by passive screen time—their language-growth trajectories can shift downward. Under such conditions, a neurotypical child may begin to follow a trajectory similar to the mild ASD curve, and a child with mild ASD may shift toward the moderate ASD trajectory. Because autistic children already experience an earlier slowing-down of syntactic-language acquisition, they are disproportionately affected by such reductions in language engagement. A neurotypical child who experiences reduced early syntactic-language engagement at home still has time to acquire the Syntactic Phenotype after entering kindergarten at age 5. In contrast, an autistic child with equally reduced syntactic engagement at home before age 5, faces a narrower developmental window, making the acquisition of the Syntactic Phenotype in later childhood significantly more challenging and, for some children, not observed ^63^.

### A simple localizational neurocognitive model

Just as a successful symphony performance depends on the precise synchronization within the orchestra, higher cognition relies on the seamless temporal coordination of distributed brain regions ^64^. This neural synchrony, in turn, rests on optimized long-range axonal connections that are largely established during early childhood as a result of engagement in syntactic conversations ^65^. A simple localizational neurocognitive hypothesis, previously proposed to account for the three distinct language phenotypes, can also explain the critical-slowing-down-ages observed in ASD and the hard boundary at puberty onset. According to the Dual Visual Stream Theory ^66^, the Ventral Stream (in the lower temporal lobe, see Figure S6) encodes the shape, size, and color of objects, whereas the Dorsal Stream (in the parietal lobe) encodes the position of objects in space. The localizational neurocognitive hypothesis linked the three language phenotypes to the developmental maturation of fronto-temporal connections to the Ventral Stream and fronto-parietal connections to the Dorsal Stream ^29^.

Within this framework, the Modifier Phenotype depends on the maturation of fronto-temporal connections, which enable the lateral prefrontal cortex (LPFC) to manipulate visual representations of object features in the Ventral Stream. This capacity supports the combination of adjectives with nouns. For example, individuals with the Modifier Phenotype can follow the instruction “*find the large green pencil”* (among pencils, straws, and Lego pieces of different colors and sizes placed on a table) by first visualizing the large green pencil in the Ventral Stream and then identifying it among other objects.

Furthermore, the Syntactic Phenotype is hypothesized to rely on mature fronto-parietal connections that enable the LPFC to combine multiple objects within the Dorsal Stream. This integrative capacity supports the comprehension of spatially relational sentences—such as those containing spatial prepositions, syntactically reversible constructions, possessive pronouns, verb tenses, and complex narratives like fairy tales. For instance, individuals with the Syntactic Phenotype can distinguish between “*the cat carries the sheep*” and “*the sheep carries the cat*” by mentally integrating the two animals and matching their internal model, encoded within the Dorsal Stream, to the correct picture.

In contrast, in the absence of mature fronto-temporal and fronto-parietal connectivity, individuals exhibit the Command Phenotype. Without these pathways, the LPFC cannot manipulate object features or integrate multiple objects within the Ventral and Dorsal Streams. Consequently, language comprehension is limited to single words and simple commands that do not require LPFC-mediated manipulation or integration. For example, the instruction “*bring the ball to Mommy*” can be executed without LPFC involvement: “*ball*” and “*Mommy*” are retrieved separately by the Wernicke’s area, and the action is implicit, as it is the only meaningful action involving these objects. While an individual with the Syntactic Phenotype could use the LPFC to mentally juxtapose the ball and Mammy, this is not necessary for understanding the command.

In neurotypical children, the critical-slowing-down-age is over four years (Figure 6). They typically reach the Command Phenotype by around 2 years of age, when they begin to follow simple commands such as “*bring the cup/ball/pencil to Mommy/Daddy/sister*” ^32^. By approximately three years of age, maturation of fronto-temporal connections enables the LPFC to exert control over the Ventral Stream, supporting the Modifier Phenotype—for example, identifying “*the small red straw*” among similar objects of varying colors and sizes. By around four years of age, further maturation of fronto-parietal tracts allows the LPFC to control the Dorsal Stream, marking the emergence of the Syntactic Phenotype, as children begin to comprehend syntactically complex explanations and narratives, such as fairy tales.

In a substantial proportion of autistic children, the critical-slowing-down-age is markedly reduced. Consequently, the rate of experience-dependent maturation of the fronto-parieto-temporal network begins to decline prematurely. Although these children could, in principle, continue to develop the fronto-parieto-temporal circuitry gradually, the onset of puberty imposes a developmental constraint possibly through synaptic pruning and cell death ^67^. Under these circumstances, even optimal language-nurturing environments, such as rich syntactic conversations and fairy tales, may be insufficient to support the emergence of the Syntactic Phenotype. Suboptimal language environments may further limit the attainable comprehension phenotype.

### Clinical and translational significance

Language-comprehension phenotype assessments hold considerable potential as both diagnostic and prognostic tools ^32^. Traditional receptive-language tests largely measure pragmatic understanding by evaluating vocabulary knowledge and word-category recognition ^68,69^. Although vocabulary size correlates with comprehension phenotype, this approach can yield misleading assessments because atypical individuals—even those with the Command Phenotype—can learn an essentially unlimited number of words ^70,71^. The limitations of assessing syntactic language by measuring vocabulary may be illustrated by the following example. Vocabulary and arithmetic skills are also strongly correlated; yet, they are recognized as distinct domains and no one evaluates arithmetic skills by measuring vocabulary. Similarly, vocabulary and syntactic-language comprehension represent different domains, and assessing one of them cannot serve as a valid proxy for assessing the other. Conversely, language-comprehension phenotype assessments target clusters of co-developing combinatorial abilities and therefore provides a more accurate measure of true language-comprehension progress.

These language-comprehension phenotype assessment instruments can enhance language therapy in several key ways:

**1. Facilitate early diagnosis of language-comprehension delay**. Failure to acquire the Modifier Phenotype by approximately 3.5 years is a strong indicator that the Syntactic Phenotype may never emerge, underscoring the need for immediate intervention. Numerous studies demonstrate that early interventions, especially language-focused exercises, significantly improve developmental outcomes ^72–77^.
**2. Prioritizing combinatorial exercises in therapy.** Language therapy for autistic children often focuses on word learning and articulation, in part because these goals are easier to target. Vocabulary training is 1) faster to administer, 2) highly valued by parents, 3) intuitive—since typically developing children accumulate large vocabularies before mastering syntax, and 4) reinforced by conventional assessments, such as Peabody Picture Vocabulary Test (PPVT-4) ^78^ and Expressive Vocabulary Test (EVT-2) ^68^. However, this strong emphasis on vocabulary can divert attention from combinatorial exercises, which are far more critical for long-term comprehension. Unlike vocabulary, which can be acquired throughout life, the Syntactic Phenotype does not develop significantly after the onset of puberty ^51,79,80^. Widespread adoption of language-comprehension phenotype assessments as benchmarks for therapeutic progress could help redirect clinical focus toward combinatorial training, enabling more children to achieve their full linguistic potential.
**3. Phenotyping may help to fine tune language therapy**. Classifying a child’s comprehension abilities into the Command, Modifier, or Syntactic Phenotypes enables targeted and developmentally appropriate intervention. For example, children who remain at the Command level may benefit most from exercises that build adjective–noun integration, whereas those at the Modifier level may be ready to begin syntactic-language training. Importantly, introducing syntactic-level exercises to a child still at the Command stage may be ineffective or even counterproductive, as it diverts valuable time away from the Modifier-level skills that the child is developmentally prepared to acquire. A finely tuned, stepwise approach ensures that therapeutic efforts align with each child’s current developmental readiness, thereby maximizing the efficiency and impact of language intervention.
**4. Facilitating clinician–parent communication**. Clinicians are often hesitant to discuss the critical period concept with parents, in part because the specifics remain unclear. Consequently, many parents delay initiating language therapy. The language-comprehension growth chart in Figure 6 can serve as a powerful visual tool for clinicians to illustrate a child’s likely language-development trajectory. By showing how learning-rates change over time—and how early intervention can meaningfully alter the outcomes—clinicians can more effectively convey the urgency of beginning therapy as early and as intensively as possible to maximize a child’s potential for acquiring the Syntactic Phenotype.
**5. Potential implications of pubertal timing for language acquisition.** Converging evidence from the present study and prior work ^49–57^ strongly indicates that puberty onset imposes a hard developmental boundary on the acquisition of the Syntactic Phenotype. Because individuals who remain at the Modifier level frequently face substantial barriers to independent living—a core concern for many families ^81^—the identification of modifiable factors that influence this trajectory is of critical clinical importance. One provocative implication of our findings is that delaying puberty onset could potentially extend the window for acquiring the Syntactic Phenotype. However, any exploration of puberty-delaying interventions would require rigorous, longitudinal clinical trials, careful ethical deliberation, and close multidisciplinary oversight to establish both efficacy and long-term safety. At present, such interventions remain purely speculative and are not endorsed for this purpose.
**6. Future potential of genetic predictors of critical slowing-down age.** In the coming years, genetic analyses may enable reliable prediction of an individual child’s critical-slowing-down-age. If validated, such predictors could identify—at a very early age—those children at greatest risk of failing to attain the Syntactic Phenotype. This knowledge would allow clinicians and families to implement intensive, evidence-based interventions during the period of peak learning-rate. Promising modifiable factors include high-dosage, developmentally tailored language therapy; substantially increased child-directed speech and joint book-reading; enriched social-play environments; strict limitation of passive screen exposure; and optimized nutritional and sleep support ^16–28^. Genetically informed risk stratification could, in principle, guide personalized intervention plans that maximally exploit each child’s biologically defined window of heightened plasticity—ultimately increasing the probability of achieving the Syntactic Phenotype.
**7. Future prospects for pharmacological extension of the critical-slowing-down-age.** In principle, novel pharmacological agents could be developed to delay the critical-slowing-down-age and thereby extend the effective duration of heightened synaptic plasticity for syntactic language acquisition. If safe and efficacious compounds emerge, they could be selectively administered to children identified by genetic or other early biomarkers as being at high risk of premature critical-slowing-down and consequent failure to reach the Syntactic Phenotype before the pubertal boundary. Such an approach would represent a transformative strategy for preventing persistent language impairment in moderate-to-severe ASD. However, any candidate intervention would first require exhaustive preclinical testing, followed by rigorously controlled, longitudinal clinical trials to establish long-term efficacy, neurodevelopmental safety, and acceptable risk–benefit profiles. At present, no pharmacological agents are known or approved for this purpose, and research in this domain remains entirely speculative.

### Limitations and future directions

This study relied on caregiver-reported assessments, which, although validated in previous large-scale datasets ^32,40,82^, may introduce subjective bias. Future work incorporating direct behavioral testing and neurophysiological measures would strengthen the link between observed phenotypes and underlying neural mechanisms. Furthermore, while the dataset spanned thousands of children, the current analysis did not systematically examine the influence of intervention type, socioeconomic status, or bilingualism, all of which may modulate progression rates.

## Conclusion

This longitudinal study demonstrates that autistic children traverse the same three discrete language-comprehension phenotypes observed in neurotypical development (Command → Modifier → Syntactic), but differ markedly in both the likelihood and timing of their acquisition, with more severe ASD associated with lower ultimate phenotype attainment and substantially delayed progression. Many autistic children remain at the Command or Modifier levels for years, a pattern most pronounced in the severe ASD. In contrast, those who do reach the Syntactic Phenotype advance rapidly, consistent with an all-or-nothing transition. The near absence of upward transitions after puberty suggests a hard maturational boundary that interacts with earlier critical-slowing-down of learning-rate in ASD, jointly constraining syntactic acquisition. Together, these findings provide the first large-scale quantification of discrete language comprehension trajectories in autism and underscore the urgent need for early, phenotype-targeted interventions during the narrow window of heightened neuroplasticity.

## Data Availability

Code and data can be downloaded from https://doi.org/10.6084/m9.figshare.30801752.

## Acknowledgements

We wish to thank all participants’ caregivers who found time to complete children’s assessments. The authors are very grateful to Dr. Petr Ilyinskii for his scrupulous editing of this manuscript. The language therapy app used to collect the data presented in this manuscript was made possible by the contributions of Rita Dunn, Alexander Faisman, Jonah Elgart, Lisa Lokshina, and Yulia Dumov. This research received no external funding.

## Author contributions

AV and EK designed the study. RV, AN, EK, and AV analyzed the data. AV and RV wrote the paper. All authors have read and approved the manuscript.

