## Supplemental material for "Longitudinal trajectories across the Command, Modifier, and Syntactic Phenotypes of language comprehension in over 6,000 autistic children"

### Supplementary Material

Supplementary Table 1: Autism Treatment Evaluation Checklist (ATEC) [1], subscale 1: Speech/Language/Communication. The answers choices were: not true, somewhat true, very true.

| |
|---|
| **1. Knows own name** |
| **2. Responds to 'No' or 'Stop'** |
| **3. Can follow some commands** |
| 4. Can use one word at a time (No!, Eat, Water, etc.) |
| 5. Can use 2 words at a time (Don't want, Go home) |
| 6. Can use 3 words at a time (Want more milk) |
| 7. Knows 10 or more words |
| 8. Can use sentences with 4 or more words |
| 9. Explains what he/she wants |
| 10. Asks meaningful questions |
| 11. Speech tends to be meaningful/relevant |
| 12. Often uses several successive sentences |
| 13. Carries on fairly good conversation |
| 14. Has normal ability to communicate for his/her age |

Supplementary Table 2: ATEC subscale 2: Sociability**.** The answers choices were: not true, somewhat true, very true.

| |
|---|
| 1. Seems to be in a shell – you cannot reach him/her |
| 2. Ignores other people |
| 3. Pays little or no attention when addressed |
| 4. Uncooperative and resistant |
| 5. No eye contact |
| 6. Prefers to be left alone |
| 7. Shows no affection |
| 8. Fails to greet parents |
| 9. Avoids contact with others |
| 10. Does not imitate |
| 11. Dislikes being held/cuddled |
| 12. Does not share or show |
| 13. Does not wave 'bye bye' |
| 14. Disagreeable/not compliant |
| 15. Temper tantrums |
| 16. Lacks friends/companions |
| 17. Rarely smiles |
| 18. Insensitive to other's feelings |
| 19. Indifferent to being liked |
| 20. Indifferent if parent(s) leave |

Supplementary Table 3: ATEC subscale 3: Sensory/Cognitive awareness**.** The answers choices were: not true, somewhat true, very true.

| |
|---|
| 1. Responds to own name |
| **2. Responds to praise** |
| 3. Looks at people and animals |
| 4. Looks at pictures (and T.V.) |
| 5. Does drawing, coloring, art |
| 6. Plays with toys appropriately |
| 7. Appropriate facial expression |
| 8. Understands stories on T.V. |
| 9. Understands explanations |
| 10. Aware of environment |
| 11. Aware of danger |
| 12. Shows imagination |
| 13. Initiates activities |
| 14. Dresses self |
| 15. Curious, interested |
| 16. Venturesome - explores |
| 17. “Tuned in” — Not spacey |
| 18. Looks where others are looking |

Supplementary Table 4: ATEC subscale 4: Health/Physical/Behavior**.** The answers choices were: not a problem, minor problem, moderate problem, and serious problem.

| |
|---|
| 1. Bed-wetting |
| 2. Wets pants/diapers |
| 3. Soils pants/diapers |
| 4. Diarrhea |
| 5. Constipation |
| 6. Sleep problems |
| 7. Eats too much/too little |
| 8. Extremely limited diet |
| 9. Hyperactive |
| 10. Lethargic |
| 11. Hits or injures self |
| 12. Hits or injures others |
| 13. Destructive |
| 14. Sound-sensitive |
| 15. Anxious/fearful |
| 16. Unhappy/crying |
| 17. Seizures |
| 18. Obsessive speech |
| 19. Rigid routines |
| 20. Shouts or screams |
| 21. Demands sameness |
| 22. Often agitated |
| 23. Not sensitive to pain |
| 24. “Hooked” or fixated on certain objects/topics |
| 25. Repetitive movements (stimming, rocking, etc.) |

Supplementary Table 5: Mental Synthesis Evaluation Checklist (MSEC) [2]. The answers choices were: not true, somewhat true, very true.

| |
|---|
| **1. Understands simple stories that are read aloud** |
| **2. Understands elaborate fairy tales that are read aloud (i.e. stories describing FANTASY creatures)** |
| 3. Draws a VARIETY of RECOGNIZABLE images (objects, people, animals, etc.) |
| 4. Can draw a NOVEL image following YOUR description (e.g. a three-headed horse) |
| 5. Engages in a VARIETY of make-believe activities (such as: playing house, playing with toy soldiers, building forts and castles, etc.) |
| **6. Understands some simple modifiers (i.e. green apple vs. red apple or big apple vs. small apple)** |
| **7. Understands several modifiers in a sentence (i.e. small green apple)** |
| **8. Understands size (can select the largest/smallest object out of a collection of objects)** |
| **9. Understands possessive pronouns (i.e. your apple vs. her apple)** |
| **10. Understands spatial prepositions (i.e. put the apple ON TOP of the box vs. INSIDE the box vs. BEHIND the box)** |
| **11. Understands verb tenses (i.e. I will eat an apple vs. I ate an apple)** |
| **12. Understands the change in meaning when the order of words is changed (i.e. understands the difference between 'a cat ate a mouse' vs. 'a mouse ate a cat')** |
| **13. Understands NUMBERS (i.e. two apples vs. three apples)** |
| 14. Can perform simple arithmetic: 2 + 3 = ? |
| 15. Can add larger numbers: 7 + 6 = ? |
| 16. Can perform simple subtraction: 3 – 2 = ? |
| 17. Can subtract larger numbers: 15 – 7 = ? |
| 18. Can perform simple multiplication: 2 × 2 = ? |
| 19. Can multiply larger numbers: 6 × 7 = ? |
| **20. Understands explanations about people, objects or situations beyond the immediate surroundings (e.g., “Mom is walking the dog,” “The snow has turned to water”)** |

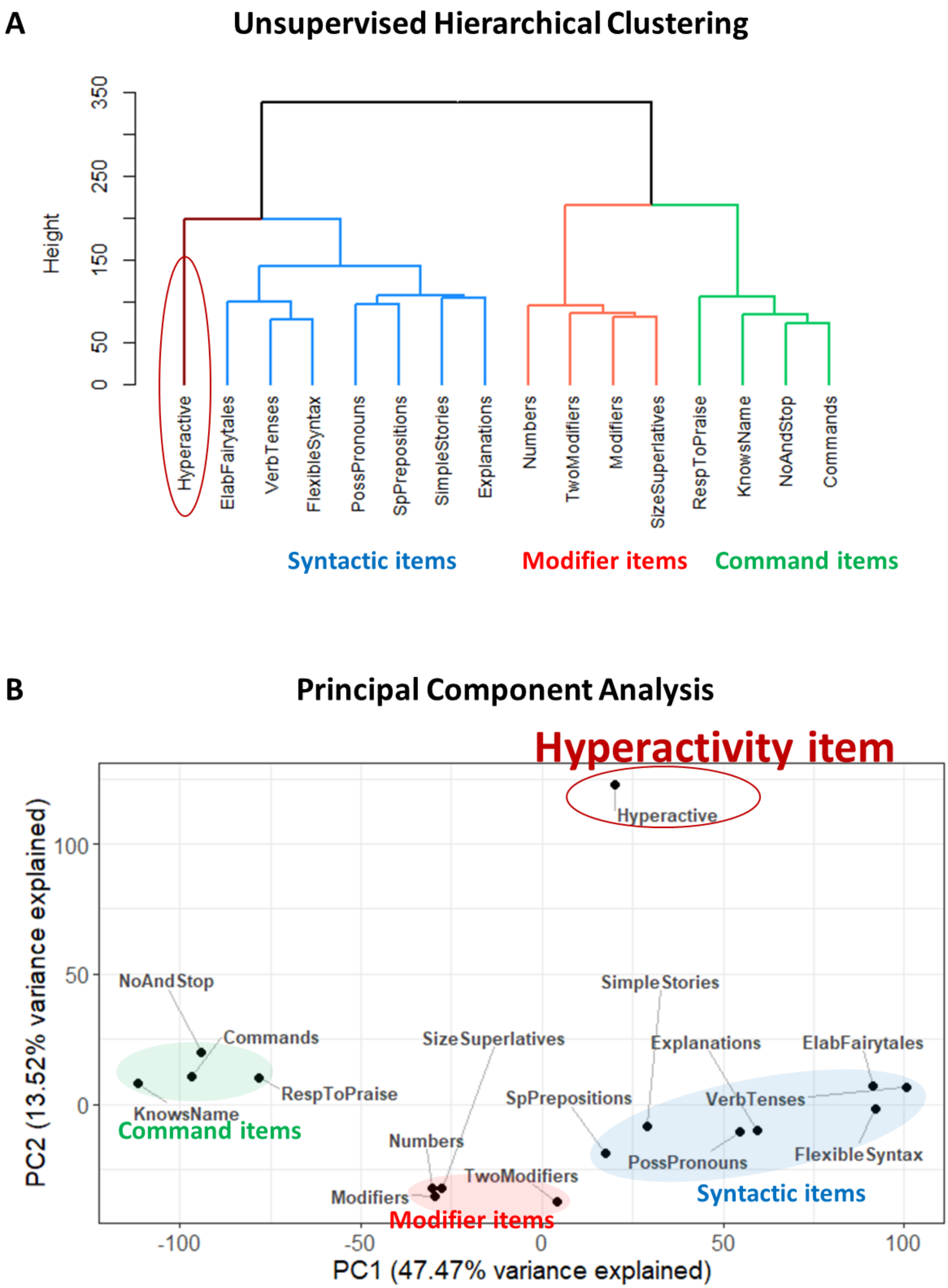


Figure S1. Clustering analysis of 15 language comprehension items with the Hyperactivity item. (A) The dendrogram representing the unsupervised hierarchical clustering of language comprehension abilities. (B) Principal component analysis.

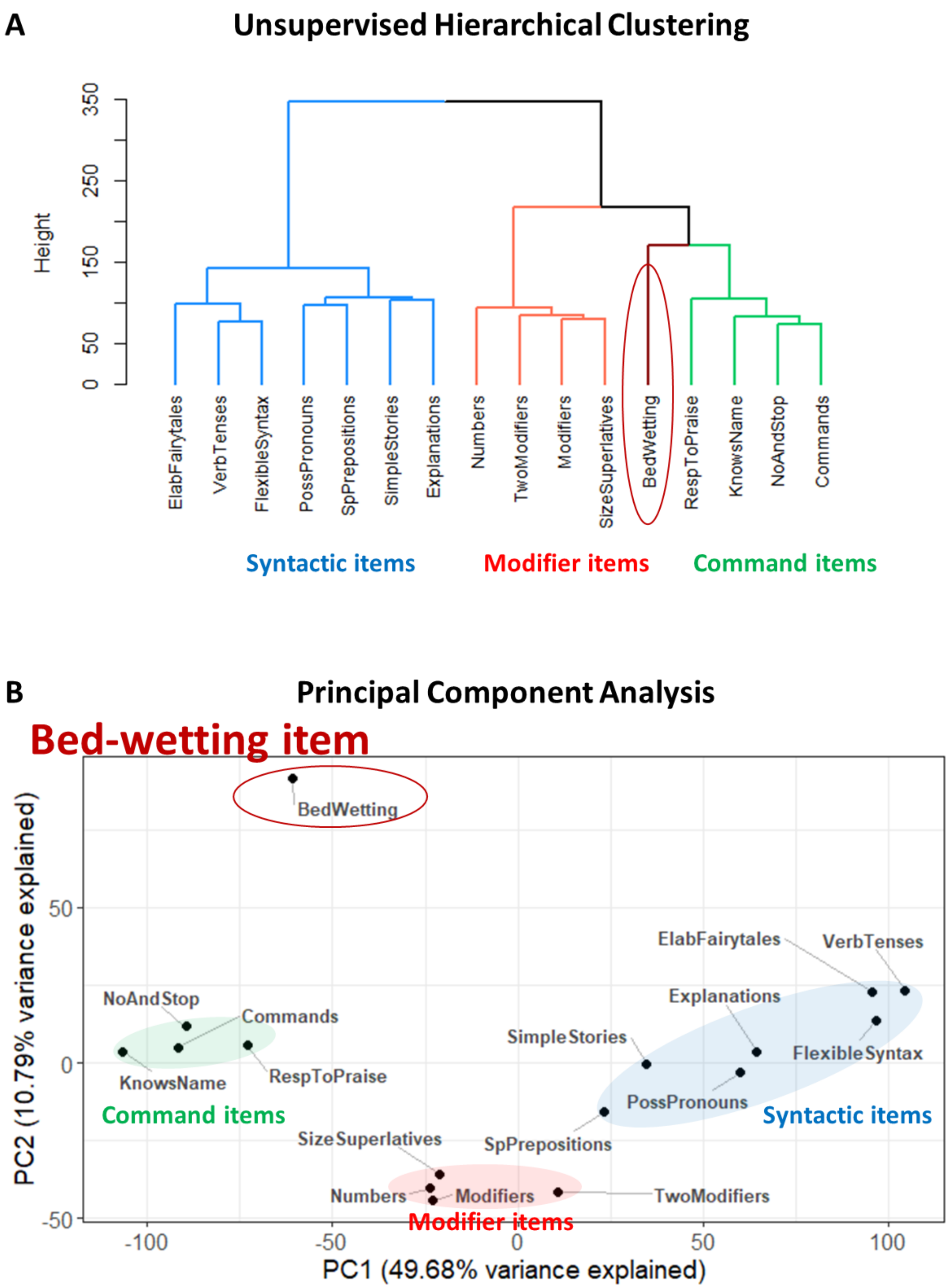


Figure S2. Clustering analysis of 15 language comprehension items with the Bedwetting item. (A) The dendrogram representing the hierarchical clustering of language comprehension abilities. (B) Principal component analysis.

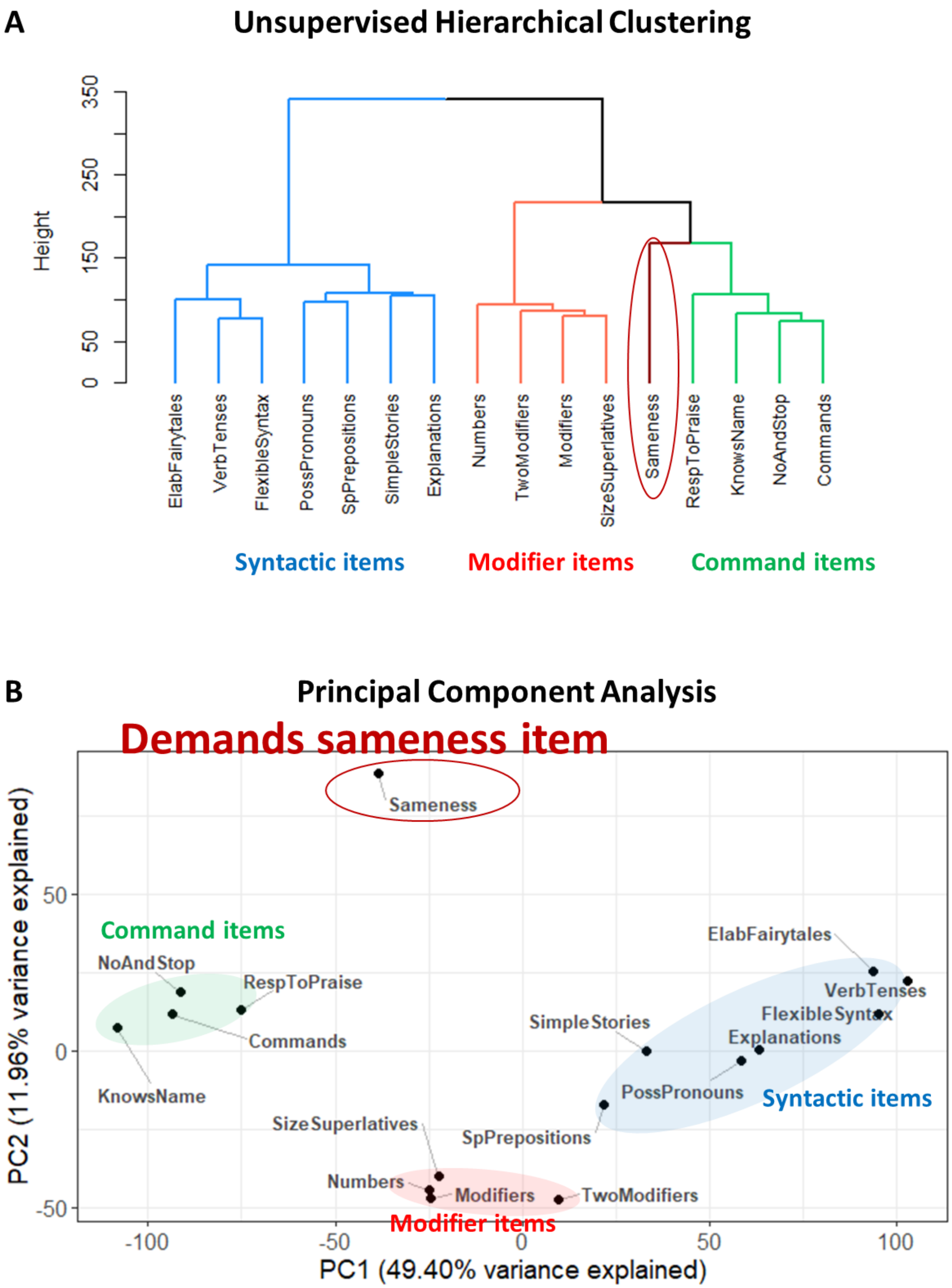


Figure S3. Clustering analysis of 15 language comprehension items with "Demands Sameness". (A) The dendrogram representing the hierarchical clustering of language comprehension abilities. (B) Principal component analysis.

Figure S4. Progression of all participants across the PCA plot as they advance through comprehension phenotypes (uploaded as mp4 and animated gif files, and also available here: https://youtu.be/MAUjiJXLwdA ). The right side of the PCA plot corresponds to the Command Phenotype, the center to the Modifier Phenotype, and the left side to the Syntactic Phenotype. Each trajectory begins at the age of the participant's first assessment and ends at the age of their last assessment, connected by a spline function computed from all available observations. The movie shows that most participants begin on the right side of the plot and gradually shift leftward over time, reflecting a developmental progression from the Command to the Modifier and, ultimately, to the Syntactic Phenotype. As in the rest of the article, neurotypical participants are shown in black, mild ASD in blue, moderate ASD in red, and severe ASD in green. Four temporal waves are discernible: 1) Dominated by neurotypical participants, peaks between 1.5 to 3.5 years of age. 2) Dominated by mild ASD, peaking between 2 to 5 years of age. 3) Dominated by moderate ASD, peaking between 2.5 to 7 years of age. 4) Dominated by severe ASD, peaking between 3 to 8 years of age. This systematic delay demonstrates that the acquisition of advanced comprehension phenotypes occurs progressively later with increasing ASD severity.

Each participant's final assessment is indicated by a circle. A prominent cloud of dark gray and blue circles appears in the leftmost region of the PCA plot, indicating that most neurotypical and mild ASD participants reach the Syntactic Phenotype. Light red circles—representing moderate ASD participants—are distributed primarily across the Modifier and Command regions. Light green circles—indicating severe ASD participants—are concentrated in the Command Phenotype region.

Leftward movement shows a pronounced decline after 5 years of age and is nearly absent after 10 years of age, indicating a substantial slowdown in progression toward higher-level phenotypes and an almost complete cessation at the onset of puberty.

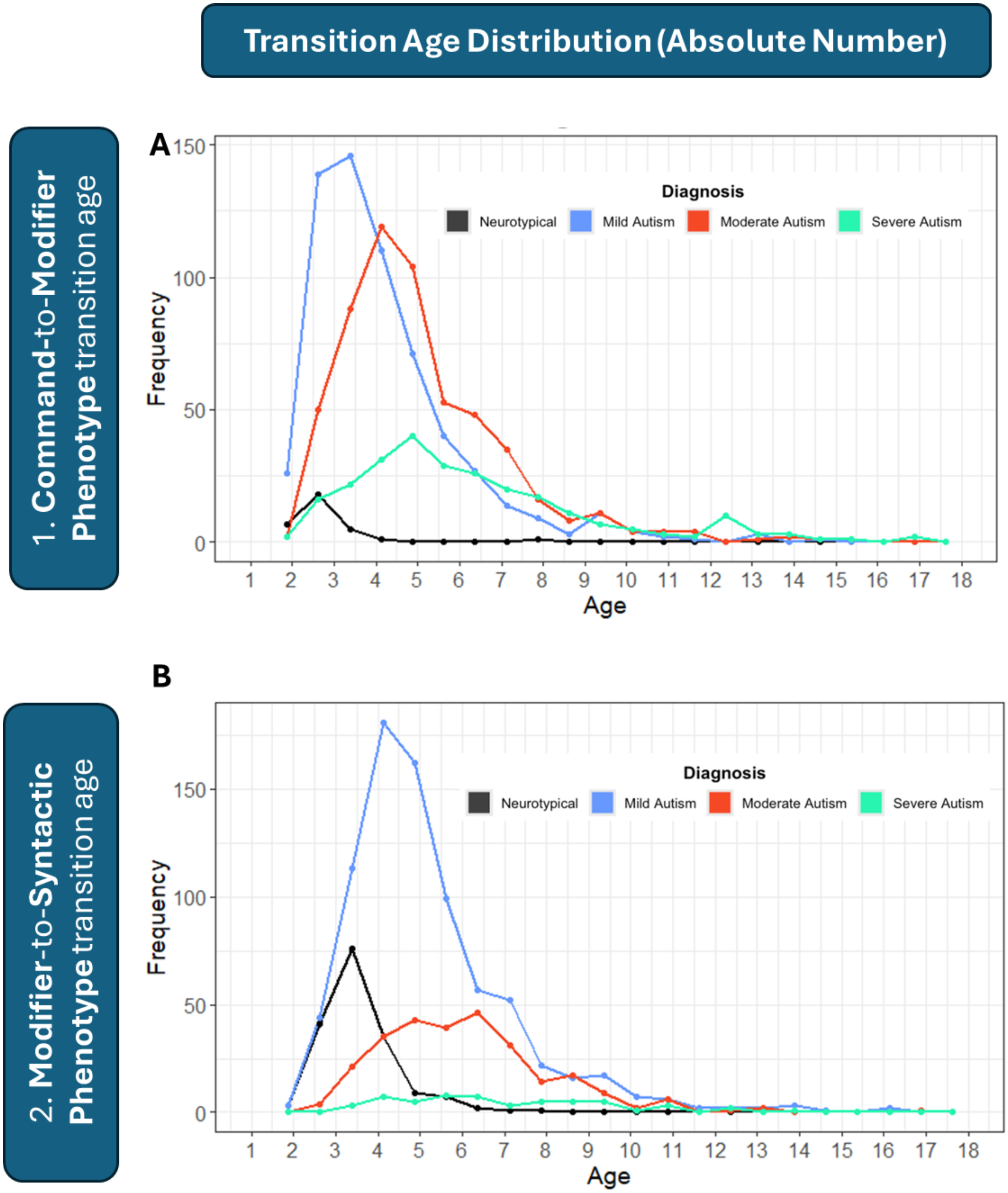


Figure 5S. Age of acquisition for each language-comprehension phenotype, shown as absolute numbers of participants. (A) Distribution of ages at which children transition from the Command to the Modifier Phenotype. (B) Distribution of ages at which children transition from the Modifier to the Syntactic Phenotype. Across all diagnostic categories, the number of participants transitioning to a higher-level phenotype declines sharply after 5 years of age and nearly completely ceases after 10 years of age.
The low number of Command-to-Modifier transitions in neurotypical children reflects the fact that most neurotypical participants already exhibit the Modifier Phenotype at baseline.
The low number of Modifier-to-Syntactic transitions in children with severe ASD reflects the fact that most individuals in this group never acquire the Syntactic Phenotype.

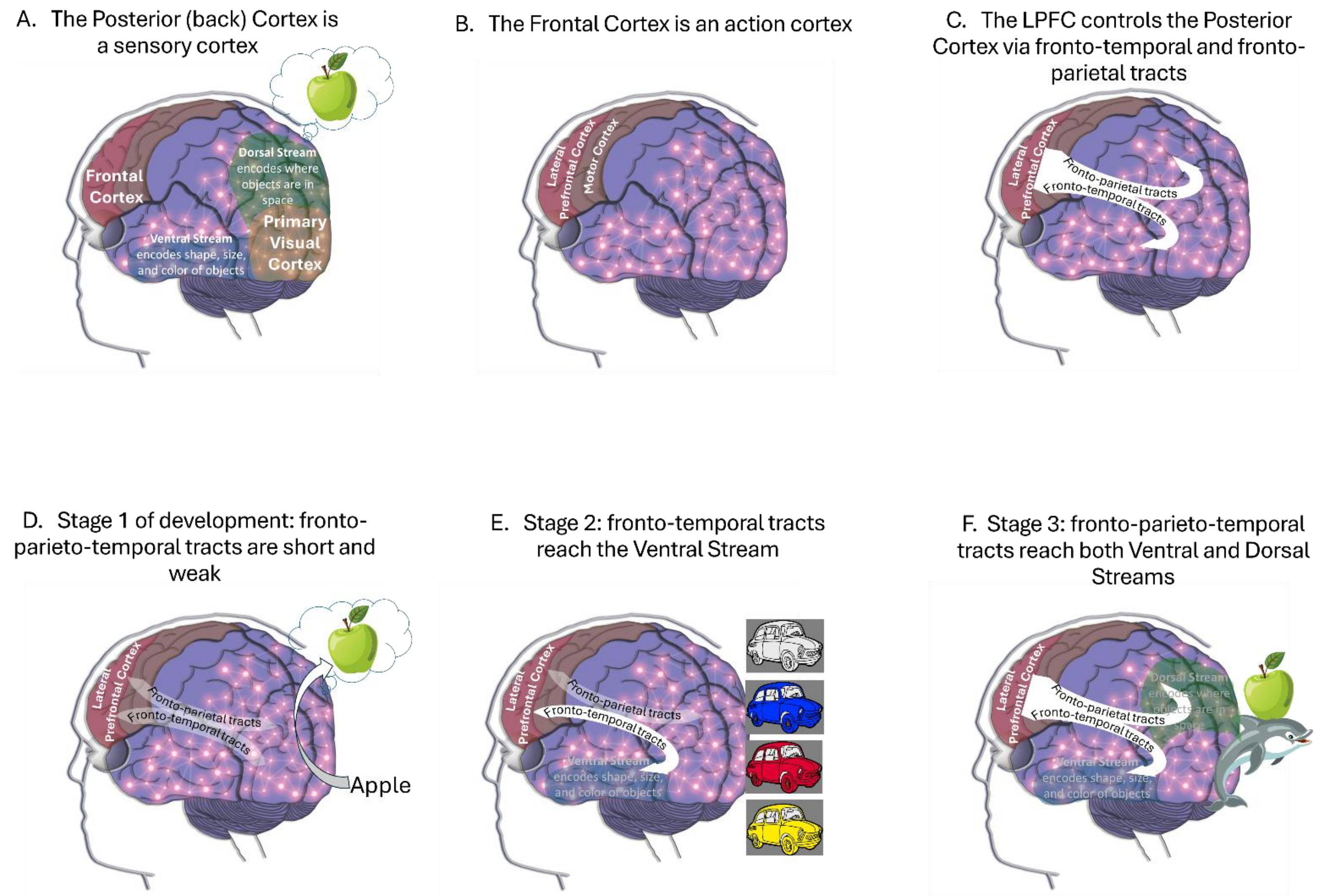


Figure 6S. A simple localizational neurocognitive hypothesis previously elaborated by Vyshedskiy et al. [3] explains the existence of the three distinct language phenotypes by examining the developmental stages of fronto-temporal and fronto-parietal connections.

(A) Sensory perceptions are encoded in the Posterior Cortex (colored in violet; the back of the brain behind the central sulcus) [4]. Visual experiences are primarily encoded in the Primary Visual Cortex, as well as the Ventral and Dorsal Streams. A group of tens of thousands of posterior cortex neurons encoding an object have enhanced synapses (illustrated by interconnected bright circles) and is called an object-encoding-neuronal-ensemble [5]. According to the Dual Visual Stream Theory [6], the Ventral Stream encodes the shape, size, and color of objects, while the Dorsal Stream encodes where objects are in space.

(B) The Frontal cortex (located in front of the central sulcus) primarily functions as the brain's action cortex [4]. The motor cortex moves muscles, while the Lateral Prefrontal Cortex (LPFC, located in front of the motor cortex) "moves" (controls) object-encoding-neuronal-ensembles in the posterior cortex.

(C) The LPFC "moves" visual objects within the mind's eye by controlling activity in the Ventral and Dorsal Streams (where those objects are encoded) via the fronto-temporal and fronto-parietal white matter tracts [7].

(D) When the fronto-parieto-temporal tracts are abnormally weak and short [8], the LPFC cannot modify an object's size or color as encoded in the Ventral Stream, nor the spatial relationships between objects encoded in the Dorsal Stream. This limitation restricts the individual to the Command Phenotype [3].

(E) When the fronto-temporal tracts reach the Ventral Stream, the LPFC control enables deliberate modification of an object's shape, size and color—a neurological mechanism termed the Prefrontal Modification (PFM) [3]. For example, an individual with PFM can deliberately imagine their car in different colors and sizes. Such an individual can combine adjectives with nouns and follow instructions like "find the small green straw" from a set of straws, Lego pieces, and pencils of various sizes and colors—by first imagining the target item and then matching the mental image to the object in the set. This individual exhibits the Modifier Phenotype.

(F) When the fronto-parieto-temporal tracts reach both the Ventral and the Dorsal Streams, the LPFC can additionally control the spatial location of objects—a neurological mechanism termed Prefrontal Synthesis (PFS) [3]. An individual with PFS can juxtapose multiple mental objects in space and simulate their interactions in the mind's eye. For example, they can imagine things that do not occur in real life, such as an apple being carried by a dolphin. Just as the premotor cortex plans muscle movements in space, the LPFC manipulates and juxtaposes objects in mental space, virtually testing their interactions. This individual can understand stories and exhibits the Syntactic Phenotype.

Deliberate mental simulations conducted by the LPFC should not be conflated with spontaneous simulations, such as REM-sleep dreams, because their underlying mechanisms differ fundamentally with respect to LPFC involvement. The LPFC remains inactive during sleep [9], and individuals with LPFC damage show no significant changes in their dreaming content [10]. In contrast, damage to the LPFC or the fronto-parieto-temporal tracts frequently impairs the PFS ability [11]. The distinction between PFS and spontaneous dreams is analogous to the difference between voluntary muscle movement controlled by the motor cortex and involuntary spasms originating in the muscle. Crucially, involuntary mental simulations, such as dreams, cannot support real-time communication—only PFS enables the rapid execution necessary for effective understanding of stories.
